# EFFICACY OF STEVIA REBAUDIANA BERTONI ON LEVELS OF GCF GLUCOSE AND BIO-MARKERS IN DIABETIC PATIENTS WITH PERIODONTITIS-A SPLIT MOUTH RANDOMIZED CONTROLLED TRIAL

**DOI:** 10.1101/2024.12.01.24318252

**Authors:** Rampalli Viswa Chandra, Nandini Komaravelli, Aileni Amarendhar Reddy

**Affiliations:** Department of Periodontics, SVS Institute of Dental Sciences, Mahabubnagar, Telangana

**Keywords:** Diabetes, Periodontitis, Gingival crevicular fluid, Stevia rebaudiana, Ghrelin

## Abstract

**Aim & Objectives:** The aim of the present study was to evaluate the effect of Stevia rebaudiana bertoni on GCF glucose levels and GCF bio-markers in diabetic patients with periodontitis.

**Patients and methods:** 19 subjects were participated in the study. Randomly, a quadrant was allotted as test site for placing Stevia gel and other quadrant was allotted as a control site for placing placebo in the gingival sulcus having PD≥ 5mm after performing thorough scaling and root planing. GCF samples was collected using sterile paper strips at baseline i.e., after scaling and root planing, 3 months and 6 months using intra-crevicular superficial method. GCF samples was eluted from the strips by placing them in Eppendorf® tubes that contained 500 micro-litre of buffer and will be stored at -80℃ until analysis was done. The levels of GCF glucose and GCF bio-markers was evaluated by using commercially available ELISA kit.

**Results:** Stevia gel was effective in decreasing the GCF glucose and increasing ghrelin levels in diabetic patients with periodontitis. There was significant difference in clinical parameters in both test and control groups.

**Conclusion:** This study concluded that application of Stevia gel in gingival crevice may be of value in controlling diabetes and periodontitis at a time.

## Introduction

Periodontitis is a widespread inflammatory disease caused by specific microorganisms or interactions between groups of specific microorganisms and response by host immune system, modified by environmental and genetic factors resulting in the loss of attachment ultimately leading to tooth loss.^1^

The inflammatory response is characterized by dysregulated secretion of host-derived mediators of inflammation such as IL-1β, IL-6, prostaglandin E2 (PGE2), TNF-α, receptor activator of nuclear factor κB ligand (RANKL) and the matrix metalloproteinases (MMPs; particularly MMP-8, MMP-9 and MMP-13), as well as T cell regulatory cytokines (e.g., IL-12, IL-18), chemokines and tissue breakdown.^2^

Pathogens of the subgingival microbiota can interact with host tissues even without direct tissue penetration and accumulate on the oral cavity to form an adherent layer of plaque with the characteristics of a biofilm. The oral cavity works as a continuous source of infectious agents and its condition often reflects progression of systemic pathologies. Periodontal infection happens to serve as a bacterial reservoir that may exacerbate systemic diseases.^2^

Diabetes mellitus is a metabolic disease usually characterized by the classic triad of polydipsia, polyuria and polyphagia; consequences of homeostasis disruption due to impaired glucose metabolism.^3^

The impaired metabolism of glucose, lipids and proteins in diabetes produces alterations in macro and micro-vascular circulation that are associated with the five classic complications of the disease i.e., retinopathy, neuropathy, nephropathy, cardiovascular complications and delayed wound-healing. Periodontal disease has been proposed as the sixth complication of diabetes, based on the highly frequent presence of both diseases in the same patient. ^4,5^

Patients with diabetes type-1 or 2 or periodontal disease show an imbalance or hyper-release of soluble cytokines against the attack of coadjuvant factors. The other risk factors include tobacco use or stress for periodontal disease and the action of viruses or toxic agents for diabetes. In response to the presence of these modifying factors, cells of both diabetic and periodontal patients release an increased amount of certain cytoactive chemicals e.g., prostaglandin E2 (PGE2), interleukin 1 (IL-1) and tumor necrosis factor-alpha (TNF-α).^6^ In this way they exhibited a two-way relation between them.

With the increase in the incidence of diabetes the need for natural non-caloric sweeteners with acceptable taste and relatively safe is exigent. Stevia, the common name for the extract stevioside from the leaves of *Stevia rebaudiana Bertoni*, is a natural, non-caloric sweetener used as a sugar substitute or as an alternative to artificial sweeteners. Stevia has been found to increase insulin sensitivity in rodent models ^7^ and to have beneficial effects on blood glucose and insulin levels in human studies,^8^ which suggests it may have a role in food intake regulation. In safety studies, no negative side effects were reported.^9^

Stevia has an anti-plaque effect because of its ability in decreasing the formation of bacterial insoluble polymers.^10^ It significantly decreases the production of TNF-α, IL-1α, slightly decreases the production of NO in stimulated cells with LPS and THP-1 showing its anti-inflammatory properties.^11^ These characteristics in combination with anti-diabetic properties could results in the effective treatment of periodontal diseases.^10^

The gingival crevicular fluid (GCF) is a serum exudate originated from the microcirculation in the gingival tissues which flows into periodontal pocket (gingival sulcus) carrying inflammatory mediators and products of tissue metabolism.^12^ Several inflammatory markers such as interleukin (IL)-1β and C-reactive protein (CRP) are present in high levels in patients with periodontitis.^13^ Since the composition of GCF somewhat mirrors the plasma, it could be used as a less invasive medium to evaluate systemic conditions including DM. However, the production of inflammatory mediators is increased locally at PDs sites.

Periodontitis and diabetes show common bio-markers like ghrelin, leptin, visfatin. The levels of bio-markers in GCF are potentially important as predictors of disease progression.^14^

Ghrelin is mainly produced in cells of the stomach; it induces appetite and thereby controls food intake and energy balance. There are 2 forms of ghrelin they are acylated ghrelin which is biologically active and deacyl ghrelin which is involved in adipogenesis and both of them have shown certain immunomodulating properties. Acylated ghrelin and to some extent deacyl ghrelin, interfere with the expression of pro-inflammatory cytokines such as tumor necrosis factor-α.^15^

The endogenous ghrelin system has over the last decade emerged as being implicated in a myriad of metabolic effects that go well beyond its initial classification as a hormone affecting food intake and growth hormone secretion. Along with ghrelin’s role in systemic metabolism, a variety of studies evaluated the therapeutic impact of ghrelin pathway modulation. While ghrelin agonism might offer potential to treat diabetic gastroparesis and anorexia associated with pathological under-weight and cachexia, ghrelin receptor antagonism might be of therapeutic value to decrease body weight under certain conditions of obesity and also to improve glucose metabolism and type 2 diabetes.^15^

This study hypothesized the biomarker levels that evaluate the state of activity of the innate immune response, the pro-inflammatory cytokine regulation and connective tissue breakdown which correlate with periodontal disease & diabetes states with increasing risk burden. The current study therefore assessed the levels of GCF biomarkers such as glucose and ghrelin before and after application of stevia gel.

## Aim & Objectives

### AIM

The aim of the study was to evaluate the efficacy of *Stevia rebaudiana bertoni* on GCF glucose levels, GCF bio-markers and measures of periodontal disease.

### OBJECTIVES

- To evaluate GCF glucose & ghrelin levels in diabetic patients with periodontitis at baseline, 3 months and 6 months before and after application of Stevia gel in gingival crevice
- To evaluate probing depth, clinical attachment level, plaque index and bleeding on probing at baseline, 3 months and 6 months before and after application of Stevia gel in gingival crevice.

## Review of Literature

### Diabetes and periodontitis

A study by **Nelson et al (1990)**^22^ concluded that periodontal disease is common in non-diabetic people, in whom most of the incident cases in this study occurred. NIDDM, however, clearly confers a substantially increased risk. Thus, periodontal disease should be considered a nonspecific complication of diabetes. The inflammatory nature of periodontal disease may hinder diabetic control and the associated oral pain and tooth loss may inhibit appropriate dietary intake.

A study by **Taylor et al (1998)**^18^ has evaluated the effects of glycemic control status on both the risk for, as well as severity of periodontal destruction over time. The temporal sequence specified in these longitudinal analyses provides evidence to support a cause-effect relationship. Subjects with poorer glycemic control had significantly greater risk for alveolar bone loss progression and the progression was more severe than in subjects without type 2 DM. Additionally, these analyses suggest that there may be a gradient in risk for any alveolar bone loss, as well as severity of progression of alveolar bone loss with poorly controlled > better controlled > no type 2 DM.

A study by **Iacopino et al (2001)**^38^ stated that periodontitis may contribute to elevated proinflammatory cytokines/serum lipids and potentially to systemic disease arising from chronic hyperlipidaemia and/or increased inflammatory mediators. These cytokines can produce an insulin resistance syndrome similar to that observed in diabetes and initiate destruction of pancreatic β cells leading to development of diabetes-induced hyperlipidaemia, immune cell alterations and diminished tissue repair capacity. It may also be possible for chronic periodontitis to induce diabetes.

A study by **Chang et al (2001)**^39^ that diabetes is a risk factor for greater periodontal destruction, whereas managing periodontitis can also contribute to better glycemic control. The underlying regulatory mechanisms are also bidirectional. The hyperglycemic status may directly alter subgingival microbial compositions, impair cellular function and change collagen metabolism. The formation of advanced glycation end-products (AGEs) can further modify the extracellular matrix and establishment of cellular receptor binding can amplify inflammation. Moreover, periodontitis also induces hyperlipidemia and insulin resistance. This cyclical relationship converges via overproduction of proinflammatory cytokines such as tumor necrosis factor-a and interleukin-1b. Thus, this study highlights the importance of maintaining periodontal health to eliminate systemic complications and meticulous metabolic control to prevent further periodontal destruction.

A study by **Lalla et al (2011)**^16^ demonstrated that diabetes mellitus is an established risk factor for periodontitis, a chronic microbially induced inflammatory disorder that affects the supporting structures of teeth. Diabetes mellitus leads to a hyperinflammatory response to the bacterial challenge in periodontitis and impairs repair; these effects are at least partly mediated by the receptor for advanced glycation end products and its ligands. Periodontitis can adversely affect glycaemic control in patients with diabetes mellitus and contribute to the development of its complications. Periodontal therapy seems to result in a modest improvement of glycaemic control in patients with diabetes mellitus. The adverse effect of periodontal infections on diabetes mellitus is potentially explained by the resulting increase in systemic inflammation, which contributes to insulin resistance.

An epidemiological study by **Preshaw et al (2012)**^17^ stated that diabetes is a significant risk factor for periodontitis and the risk of periodontitis is greater if glycaemic control is poor; people with poorly controlled diabetes are at an increased risk of periodontitis and alveolar bone loss. Controlling diabetes (i.e., improving glycaemic control) is likely to reduce the risk and severity of periodontitis.

A study by **Jimenez et al (2012)**^20^ provides further support for diabetes as a risk factor for periodontitis. This is among the largest prospective studies to evaluate this association with comprehensive control of confounders. An association between diabetes and periodontitis may potentially affect a large proportion of the population. Moreover, individuals with diabetes may also be at an increased risk of tooth loss due to increased risk of periodontitis. These results hold important public health implications due to the associations between periodontitis and cardiovascular disease and nutritional alterations associated with tooth loss. Greater collaboration between diabetes care providers and dentists could be used to identify at risk patients in both clinical settings.

A study by **Morita et al (2012)**^21^ stated that the risk of developing periodontal disease was associated with levels of HbA1c and the risk of elevations of the level of HbA1c was associated with having periodontal pockets of more than 4 mm. These findings suggest that the level of HbA1c and progress of periodontal disease have a bi-directional relationship.

A study by **Chiu et al (2015)**^19^ stated that a significant bidirectional relationship was found between hyperglycaemia and PD, suggesting that both diseases may share common latent traits and pathways.

### Stevia and diabetes

A study by **Gregersen et al (2004)**^32^ concluded that stevioside reduces postprandial blood glucose levels in type 2 diabetic patients, indicating beneficial effects on the glucose metabolism. Stevioside may be advantageous in the treatment of type 2 diabetes.

A study by **McAllister et al (2007)**^23^ suggested that natural alternative sweeteners may reduce hyperglycemia, improve lipid metabolism and have antioxidant effects particularly in those that have baseline diabetes. Diabetes and metabolic syndrome have become a global healthcare crisis and the sugar overconsumption plays a major role. The use of artificial sweeteners has become more prevalent to improve insulin resistance in those with diabetes, obesity and metabolic syndrome, although the evidence does not support this result. Furthermore, it suggests that natural alternative sweeteners may be a better alternative to sugar and artificial sweeteners.

A study by **Goyal et al (2010)**^26^ stated that Stevia is safe for diabetics, as it does not affect blood sugar levels. Stevia does not have the neurological or renal side effects as other artificial sweeteners. Stevia possesses anti-fungal and anti-bacterial properties in addition to its other versatile uses. It can be safely used in herbal medicines, tonics for diabetic patients and also in daily usage products such as mouthwashes and toothpastes.

A study by **Shivanna et al (2012)**^29^ suggested that Stevia leaves do have a significant role in alleviating liver and kidney damage in the STZ-diabetic rats besides its hypoglycemic effect. It might be adequate to conclude that Stevia leaves could protect rats against streptozotocin induced diabetes, reduce the risk of oxidative stress and ameliorate liver and kidney damage.

A study by **Akbarzadeh et al (2015)**^30^ concluded that prescription of Stevia in a dose of 250 and 500 mg/d decreases the omentin level indirectly via activating insulin sensitivity and lowering blood glucose in STZ-induced diabetic rats.

A study by **Carrera et al (2017)**^28^ stated that these glycosides and the extracts from SR have pharmacological and therapeutic properties including antioxidant, antimicrobial, antihypertensive, antidiabetic and anticancer. This work reviews the antiobesity, antihyperglycemic, antihypertensive and antihyperlipidemic effects of the majority of glycosides and aqueous/alcoholic extracts from the leaves, flowers and roots of the SR. These compounds can serve as a natural and alternative treatment for diseases that are associated with metabolic syndrome, thus contributing to health promotion.

A study by **Llic et al (2017)**^31^ stated that low dose of stevioside might have some anti-diabetic properties. Consequently, there might also exist a possibility for potential use of low dose stevioside in prevention and treatment of diabetes.

A study by **Samuel et al (2018)**^24^ stated that Stevia leaf extract sweeteners are a beneficial and critical tool in sugar and calorie reduction, diabetes, weight management, and healthy lifestyles. Recent innovations have resulted in better tasting, natural-origin, high-purity Stevia leaf extracts that help both product developers and consumers make the switch from full-calorie/sugar products to reduced or zero-calorie/sugar added products to assist in meeting dietary guidelines consistent with current health and nutrition policy recommendations.

A study by **Rojas et al (2018)**^27^ stated that Stevia has potential benefits against inflammation, carcinogenesis, atherosclerosis glucose control and hypertension. On the other hand, the growing popularity of artificial sweeteners does not correlate with improved trends in obesity. An increased intake of artificial non-caloric sweeteners may not be associated with decreased intake of traditional sugar-sweetened beverages and foods. The effects of Stevia on weight change have been linked to bacteria in the intestinal microbiome, mainly by affecting Clostridium and Bacteroides species populations. A growing body of evidence indicates that Stevia rebaudiana Bertoni is protective against malignant conversion by inhibition of DNA replication in human cancer cell growth in vitro.

A study by **Farhat et al (2019)**^25^ suggested that Stevia has at least a neutral effect on short-term food intake (it did not increase food palatability) and its consumption led to lower postprandial glucose levels compared to sucrose, providing more evidence that the link between Type 2 diabetes, obesity and the consumption of NNS (non-nutritive sweeteners) is due to reverse causality.

A study by **Ajami et al (2020)**^33^ showed that the highlighted doses of Stevia in sweetened tea could be an alternative to sucralose in diabetic patients with no effects on blood glucose, HbA1C, insulin and lipid levels.

A study by **Ray et al (2020)**^35^ stated that Stevia has favorable effects on glucose homeostasis by increasing glucose-mediated insulin secretion while decreasing gluconeogenesis. That is without causing hypoglycemia. It can potentially induce weight loss and is associated with decreased inflammatory markers such as IL-6 and TNF-α, as well as oxidized-LDL and decreased atherosclerosis.

A study by **Jan et al (2021)**^34^ concluded that Stevia is full of many important phytochemicals (Steviol, Stevioside, rebaudiosides, etc.) that have properties to reduce blood sugar levels. It possesses high anti-hyperglycemic activity and serves as a substituent for saccharose in diabetes patients. It has beneficial activities in pancreatic tissue by increasing the insulin level and enhances antidiabetic properties. It also helps in maintaining normal blood sugar level by lowering inflammation and oxidative response.

### Stevia other uses

A study by **Boonkaewwan et al (2006)**^37^ stevioside induces TNF-R, IL-1a, and NO production in unstimulated human monocytic THP-1 cells. Therefore, consumption of stevioside may also be able to enhance innate immunity and protect against inflammatory diseases.

A study by **Subramaniam et al (2011)**^36^ stated that antidiabetic property of stevioside coupled with demonstrated antimicrobial property can lead to cheap and readily available natural drugs for the control of diabetes, periodontal disease and candidiasis.

A narrative review by **Contreras et al (2013)**^10^ concluded that Stevia as a potential complement in odonatological care, especially in patients that present base conditions such as obesity, diabetes and high blood pressure.

A study by **Devanoorkar et al (2014)**^43^ suggested that the levels of resistin are increased in the patients with chronic periodontitis compared to the clinically healthy controls. Resistin induces insulin resistance and large amount of resistin is secreted by adipocytes. Increased resistin levels in periodontitis may thus be considered to pose a risk for diabetes by decreasing the insulin sensitivity. Thus, periodontitis might lead to development of type II diabetes or diabetes might influence the occurrence or progression of periodontitis. Identification of resistin may lead to a specific and sensitive diagnosis.

A study by **Mohamed et al (2015)**^14^ concluded that chronic periodontitis adversely influences the GCF levels of glucoregulatory bio-markers, as it is associated with disturbed levels of bio-markers related to the onset of type 2 diabetes mellitus and its complications.

A study by **Jentsch et al (2017)**^15^ concluded that levels of ghrelin in gingival crevicular fluid are low which might be linked to periodontitis and obesity or overweight.

**Vandana et al (2017)**^11^ study concluded that Stevia demonstrated very potent at the end of 6 months trial.

### GCF and Biomarkers

A study by **Bagchi et al (2014)**^43^ suggested that the levels of resistin are increased in the patients with chronic periodontitis compared to the clinically healthy controls. Resistin induces insulin resistance and large amount of resistin is secreted by adipocytes. Increased resistin levels in periodontitis may thus be considered to pose a risk for diabetes by decreasing the insulin sensitivity. Thus, periodontitis might lead to development of type II diabetes or diabetes might influence the occurrence or progression of periodontitis. Identification of resistin may lead to a specific and sensitive diagnosis.

**Yilmaz et al (2014)**^40^ assessed the chronic periodontitis patients to determine the plasma levels of ghrelin. The author evaluated 35 chronic periodontitis & systemically healthy individuals. The parameters used for correlation were serum cytokines and bone turnover markers. The author concluded that no significant correlation was found in ghrelin levels and periodontal parameters.

The results of this study by **Mohamed et al (2015)**^14^ demonstrate that chronic periodontitis adversely influences the GCF levels of glucoregulatory biomarkers, as it is associated with disturbed levels of biomarkers related to the onset of T2DM and its medical complications.

A study by **Patnaik et al (2015)**^41^ chemerin can be considered as a potential biomarker of periodontal disease. The highest levels of the inflammatory mediator in type-2 DM with CP may indicate active inflammatory process locally in GCF and tear fluid and thus suggesting a correlation between these two secretory fluids in conditions such as diabetes and periodontitis. These biomarkers can thus be valuable in detecting high-risk individuals with periodontitis and systemic diseases like diabetes.

**Dogan et al (2016)**^42^ conducted the study to estimate the levels of chemerin and IL-6 levels. The author concluded that periodontitis and type 2 diabetes mellitus increase the release of levels of chemerin. Conversely, non-surgical therapy reduces the chemerin levels in chronic periodontitis with and without type 2 diabetes mellitus.

A study by **Bozkurt et al (2016)**^48^ stated that the levels of vaspin and omentin-1 in GCF were correlated with periodontal disease progression and hyperglycaemic condition.

A study by **Devanoorkar et al (2016)**^43^ showed that periodontitis and T2DM induced aberrant secretion of chemerin and non-surgical periodontal therapy had influence on the decrease of GCF chemerin levels both in CP patients with and without T2DM. Furthermore, it suggested that GCF chemerin level may be considered as a potential proinflammatory marker for diabetes, periodontal disease and treatment outcome.

**Jentsch et al (2017)**^15^ evaluated the levels of ghrelin and chemerin in relation with dietary update and obesity in periodontally healthy and diseased patients who differs in their body mass. The author reported that the amount of ghrelin found in GCF was lower in chronic periodontitis and obese subjects. The author concluded that lesser ghrelin and higher chemerin levels in the GCF might be co-related with the periodontal disease and obese individuals.

A study by **Yilmaz et al (2018)**^44^ stated that both diabetes and periodontitis affect hBD-1 levels in GCF.

A study by **Ahuja et al (2019)**^46^ concluded that in patients with Chronic periodontitis and Chronic periodontitis with T2DM, there might be an increase in serum leptin levels and decrease in GCF leptin levels with an increase in the severity of periodontal destruction, which could further aggravate the periodontal destruction. NSPT improved glycaemic control in patients with CP with T2DM, and also decreased the circulating levels of serum leptin and increased GCF leptin levels suggesting the possible role of leptin in mediating immune response.

A study by **Bajaj et al (2020)**^47^ concluded that Ghrelin levels shown to have a positive relation with osteoprotegerin levels and negatively relation with the levels of secretory RANKL whereas Chemerin binds to the receptors and modulates the immune system, chemotaxis of immature dendritic cells & macrophages. Further, chemerin leads to attract ChemR 23 which exposes the defence cells to adhesion molecules and extracellular matrix proteins. Thus, periodontitis can play possible role in circulating ghrelin and chemerin levels and they can be used as a marker of inflammation in periodontal diseases.

A study by **Joshi et al (2021)**^45^ stated that GCF resistin concentration was found to be significantly higher in diabetic individuals with periodontitis as compared to healthy subjects with gingivitis at baseline. There was no correlation found between HbA1c value and GCF resistin concentration, signifying the fact that increased resistin is more so related to the inflammatory condition rather than the glycaemic state of the individual.

## Patients & Methods

### STUDY DESIGN

The current study was devised as split mouth randomized control trial and included diabetic subjects with periodontitis. According to the chosen criteria, a total of 19 subjects were recruited belonging to both the genders with the age range of 20 to 70 years.

The study was conducted in the Department of Periodontics, SVS Institute of Dental Sciences, Appanapally, Mahbubnagar from September 2019 to August 2020. The study was approved by the ethical committee and scientific committee of the institutional review board with the approval number: **SVSIDS/perio/3/2019**. The study was also registered in the clinical trials registry with the registration number: **NCT04381598**

All participants willing to take part in the study were explained on the purpose and procedures of the study after which written informed consent was obtained. (Annexure I & II)

### SAMPLE SIZE CALCULATION

A sample size of 19 per group was calculated (total 38) for an effect size of 0.7, probability of α error of 0.05 and a desired statistical power of 0.8.

### SELECTION CRITERIA

#### A) INCLUSION CRITERIA

a. Subjects included in the study were in the age group ranging from 20-70 years with established diabetes and under medication to maintain normal glycaemic state i.e., Hb1Ac< 7%.
b. Oral findings included the presence of minimum of 16 natural teeth with minimum of 4 nonadjacent sites showing pocket depths ≥ 5mm.

#### B) EXCLUSION CRITERIA

Subjects who used any mouth-rinses or underwent any periodontal therapy and who took antibiotics within the 3 months were excluded.

### STUDY GROUPS

Based on the selection criteria, a total of 19 patients were enrolled from the Department of Periodontics, SVS Institute of Dental College, Mahbubnagar and a split mouth randomized control study was conducted where one side was considered as test site and other as control site.

### CONSORT FLOW CHART OF THE STUDY

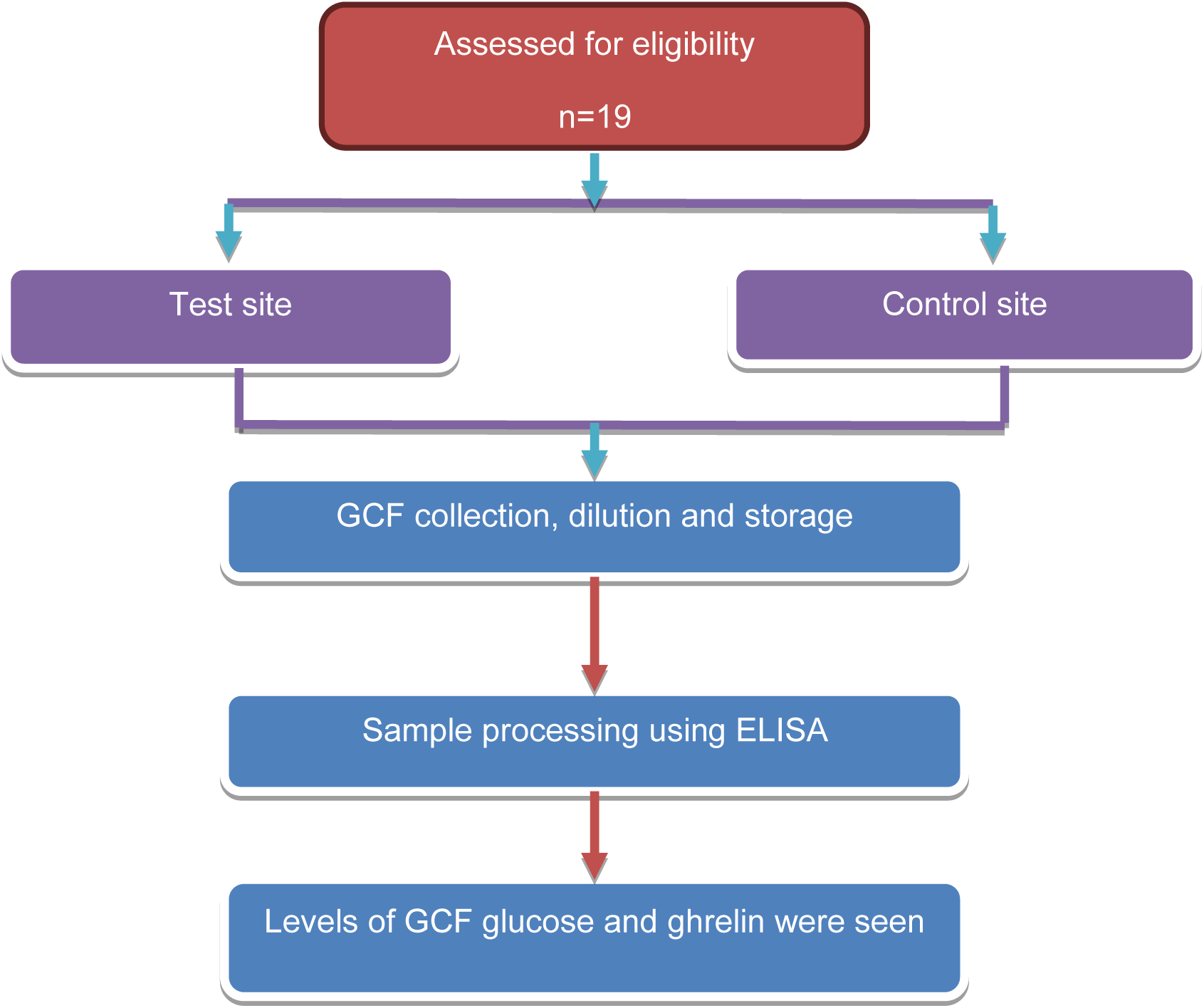

### CLINICAL EXAMINATION

A full mouth periodontal examination was carried out and the following clinical parameters were evaluated prior to sample collection by single examiner.

A. Turesky modification of Quigley-Hein index
B. Probing pocket depth
C. Clinical attachment level
D. Modified sulcular bleeding index (Mombelli et al 1987)

### PLAQUE INDEX (Turesky modification of Quigley-Hein index)

The plaque assessment was made on 6 sites per tooth (mid-buccal, mid-palatal/lingual, mesio-buccal, disto-buccal, mesio-palatal/lingual and disto-palatal/lingual) using mouth mirror and explorer. The sites were scored according to the following criteria.

**Table 1:**
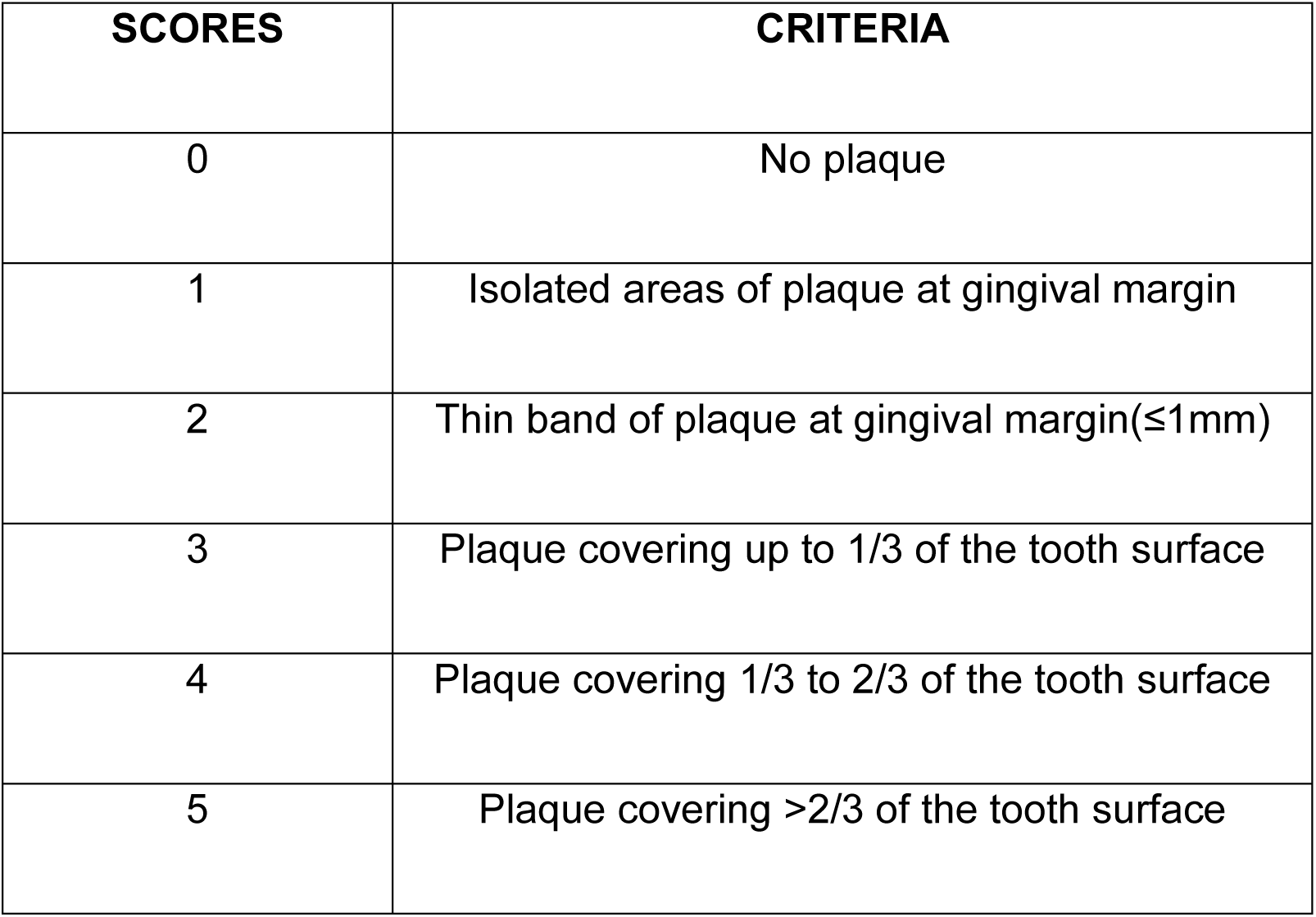
Scoring criteria of Plaque Index Calculation: PI= Total scores /No. of sites examined

**Table 2:** INTERPRETATION

| RATING | SCORE |
| --- | --- |
| Good | 0.1-0.9 |
| Fair | 1.0-1.9 |
| Poor | 2.0-3.0 |

### PROBING POCKET DEPTH EVALUATION

Probing depth was noted to be the distance from free gingival margin to the base of the gingival sulcus at 6 sites per tooth (mid-buccal, mid-palatal/lingual, mesio-buccal, disto-buccal, mesio-palatal/lingual and disto-palatal/lingual) using mouth mirror and UNC-15 periodontal probe. The readings were rounded off to the nearest millimeter.

### CLINICAL ATTACHMENT LEVEL

Clinical attachment level was assessed as the distance between cemento-enamel junction and the base of the gingival sulcus at six sites per tooth similar to probing depth using UNC-15 probe. The readings were rounded off to the nearest millimeter.

#### MODIFIED SULCULAR BLEEDING INDEX (Mombelli et al 1987)

To record the bleeding on probing values, the modified sulcular bleeding index was used. Each tooth was given a score based on the following criteria.

**Table 3:** Scoring criteria of mSBI Calculation: mSBI= Total scores/No. of teeth examined

| **SCORE** | **DESCRIPTION** |
| --- | --- |
| **0** | No bleeding when a periodontal probe is passed along the gingival margin. |
| **1** | Isolated bleeding spots visible. |
| **2** | Blood forms a confluent red line on margin. |
| **3** | Heavy or profuse bleeding |

### RADIGRAPHIC EXAMINATION

Bone loss was assessed radiographically using orthopantomogram (OPG) which aided in confirming the diagnosis of Periodontitis.

### LABORATORY EXAMINATION

All subjects were tested for post-prandial glucose levels prior to sample collection. For values nearing the diabetic range and for individuals with diabetic history, the glycemic status was further assessed using HbA1c test measuring the blood levels of glycated hemoglobin and the results were interpreted according to the American Diabetes Association guidelines. Patients with Hb1Ac levels ≤ 7% were included in this study.

**Table 4:**
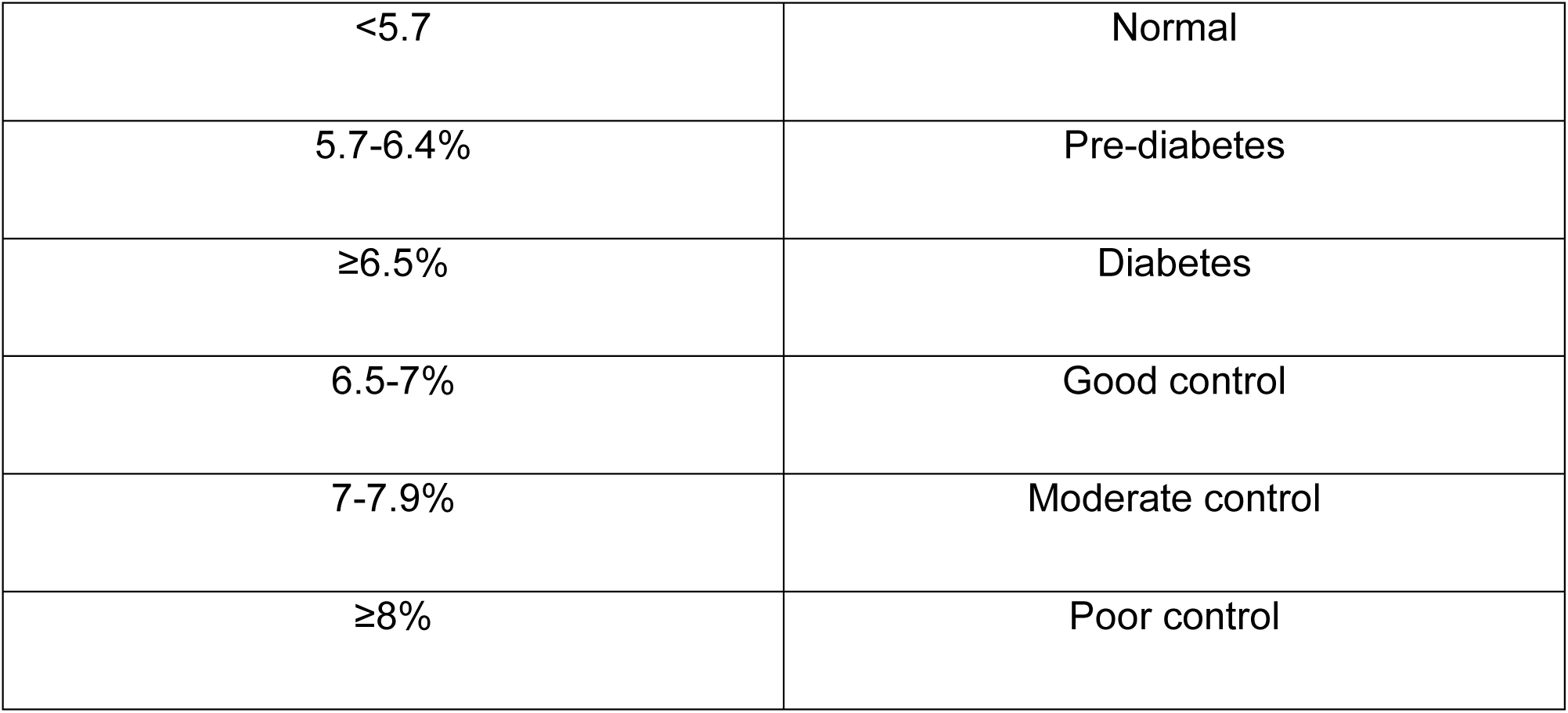
Interpretation of Glycosylated Hemoglobin:

### PROCEDURE

Randomly, a quadrant was allotted as test site for placing Stevia gel and other quadrant as control site for placing placebo in the gingival sulcus having PD≥ 5mm after performing thorough scaling and root planing. GCF samples was collected using sterile paper strips at baseline i.e., after scaling and root planing, 3 months and 6 months using intra-crevicular superficial method. GCF samples was eluted from the strips by placing them in Eppendorf® tubes that contained 500 micro-liter of buffer and was stored at -80℃ until analysis was done. The levels of GCF glucose and GCF bio-markers was evaluated by using commercially available ELISA kit.

#### Collection of GCF sample

Gingival crevicular fluid samples were collected under relative dryness using cotton rolls. At the deepest site of each quadrant in patients with periodontitis or at the mesio-buccal sites of first molars in individuals without periodontitis, sterile paper strips were placed in the periodontal pocket or gingival sulcus for 30 seconds. Then, the strips were pooled and placed in a tube containing 1 mL of 0.9% (w/v) sodium chloride.

### PRINCIPLE OF ELISA

ELISA is an assay technique used for detection and quantification of proteins, antibodies and hormones. It involves bonding between antigen and antibody to form an immune-complex that is disclosed through an enzymatically derived colorimetric reaction. The concentration of the antigen decides the color and the results are read photometrically for optimal quantification.

### ENZYME LINKED IMMUNOSORBENT ASSAY FOR GCF GLUCOSE AND BIOMARKER LEVELS

The levels of ghrelin and glucose were measured using commercially available kits (Ghrelin: Ghrelin Human ELISA Kit, Invitrogen, Mumbai, India; Glucose: Glucose Colorimetric Detection Kit, Thermo Fisher Scientific, Hyderabad, India). The assays were executed according to the manufacturer’s instructions.

#### Statistical analysis

All the variables in the study were subjected to statistical analysis using SPSS software package applying T test and ANOVA.

**Fig no.1.**
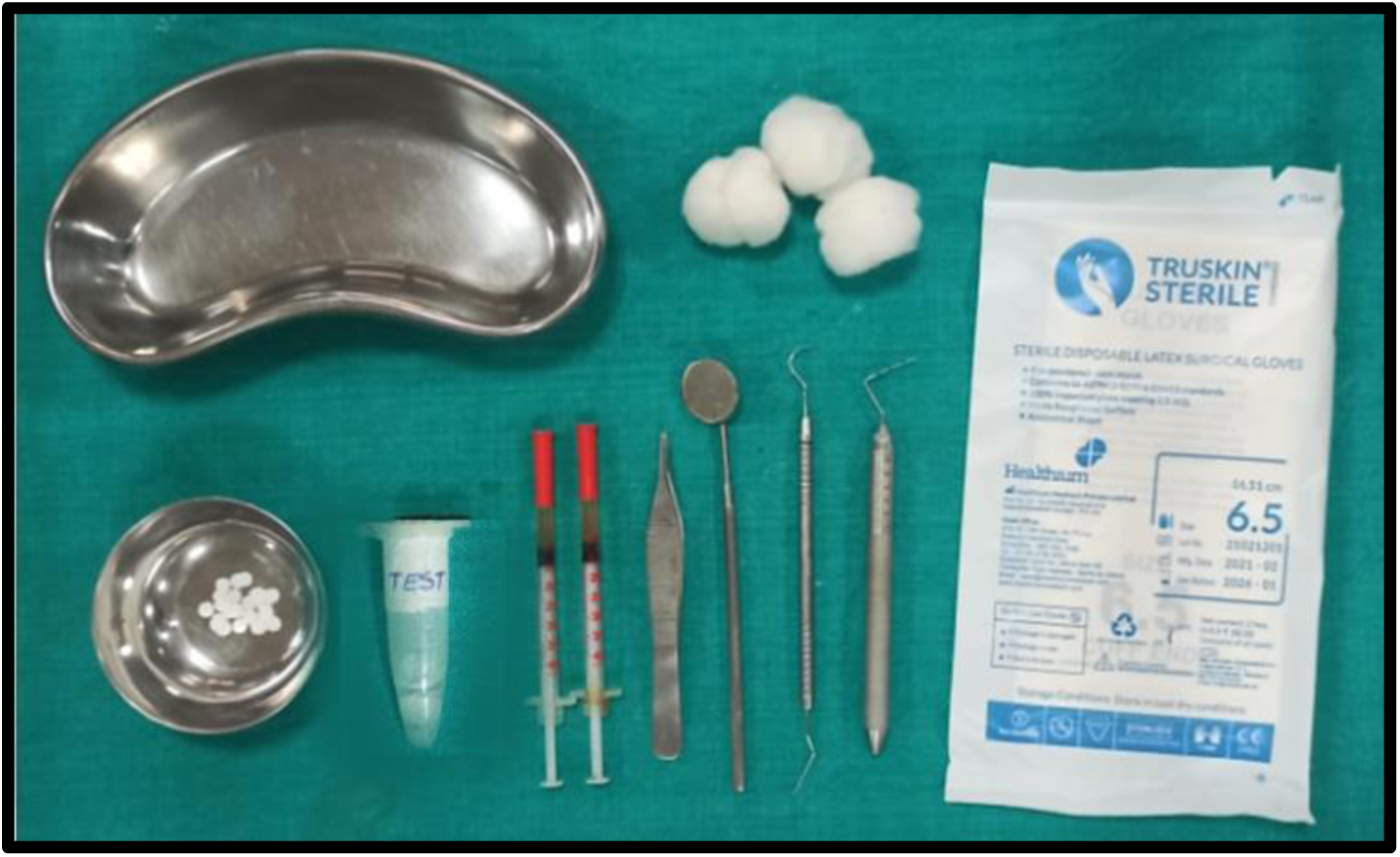
Armentarium used for GCF collection

**Fig no.2.**
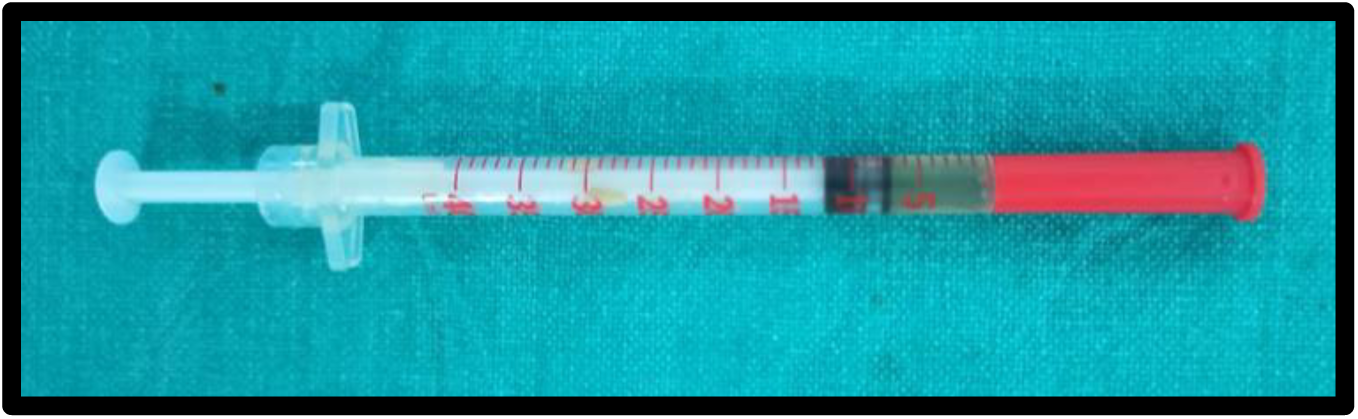
Stevia gel

**Fig no.3.**
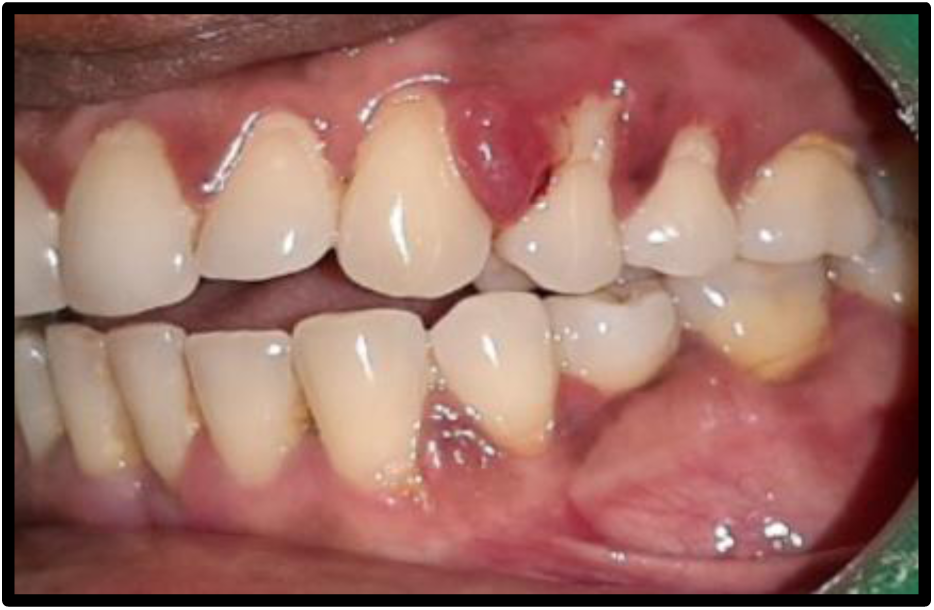
Preoperative image of test site

**Fig no.4.**
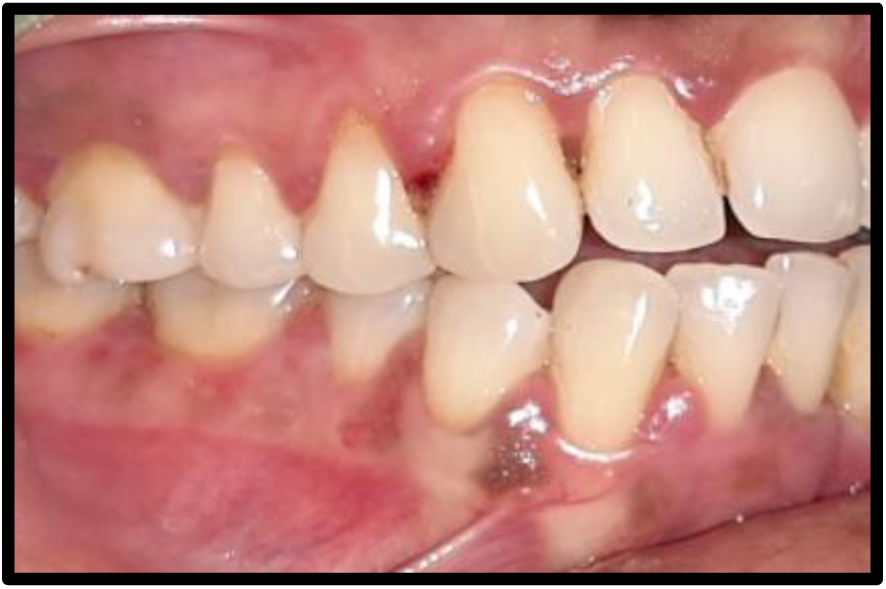
Preoperative image of control site

**Fig no.5.**
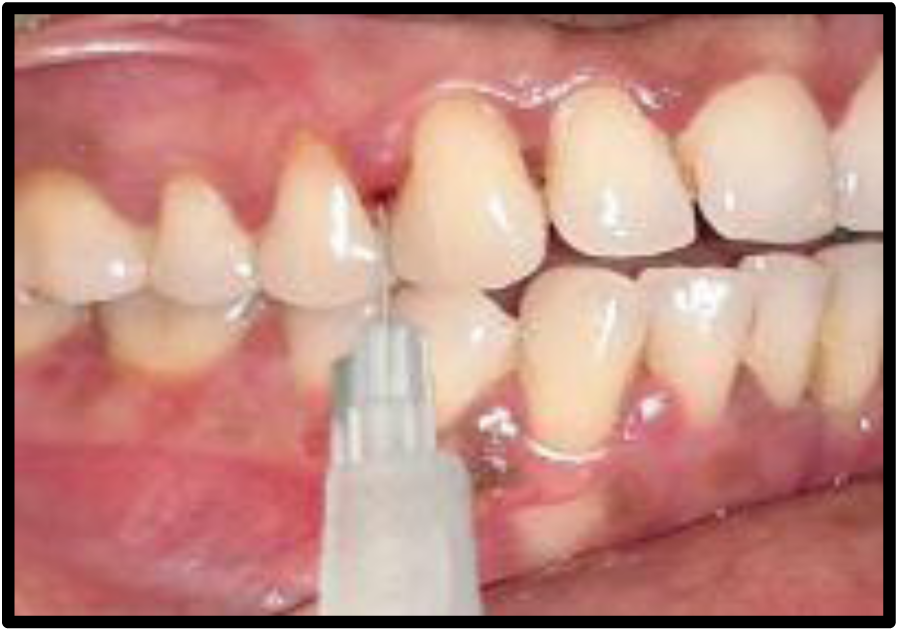
Placebo placed at control site

**Fig no.6.**
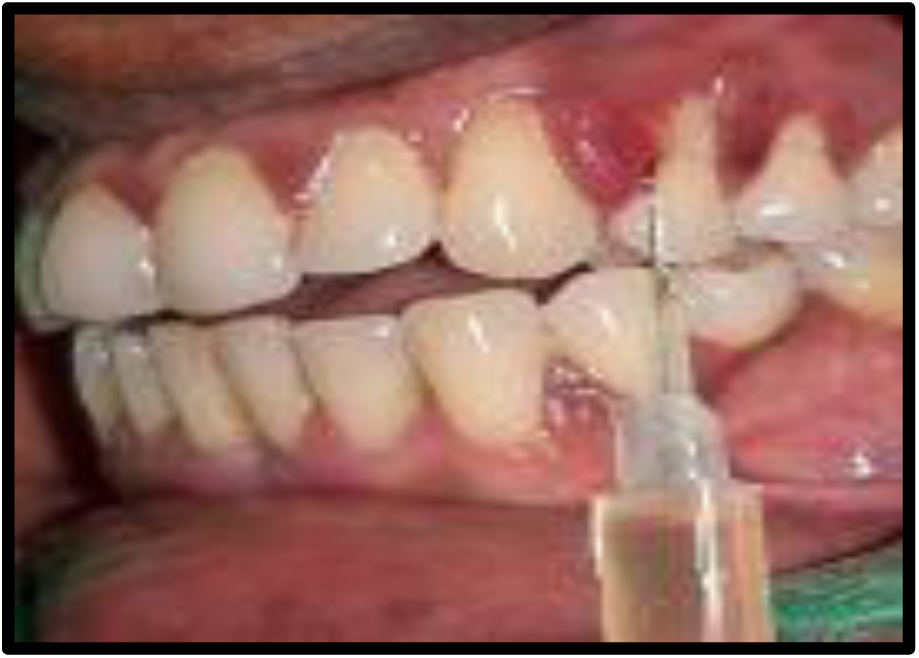
Stevia gel placed at test site

**Fig no.7.**
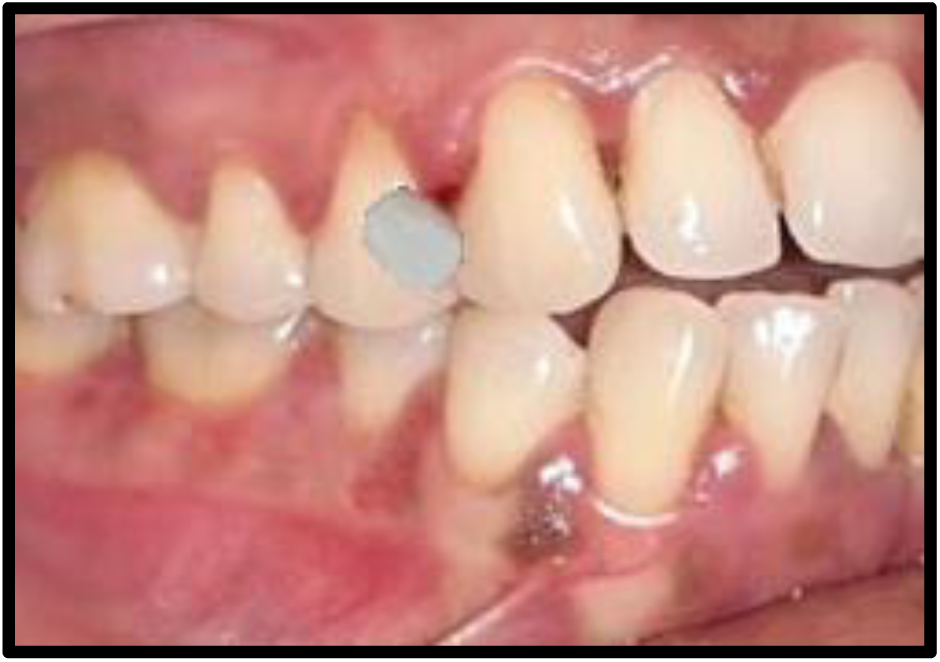
GCF collection at control site

**Fig no.8.**
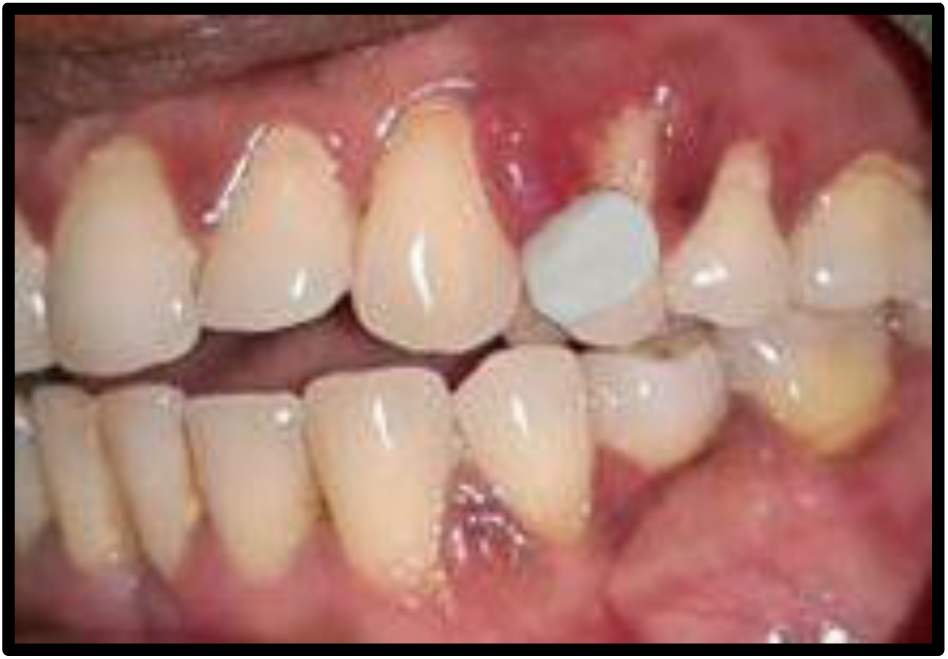
GCF collection at test site

**Fig no.9.**
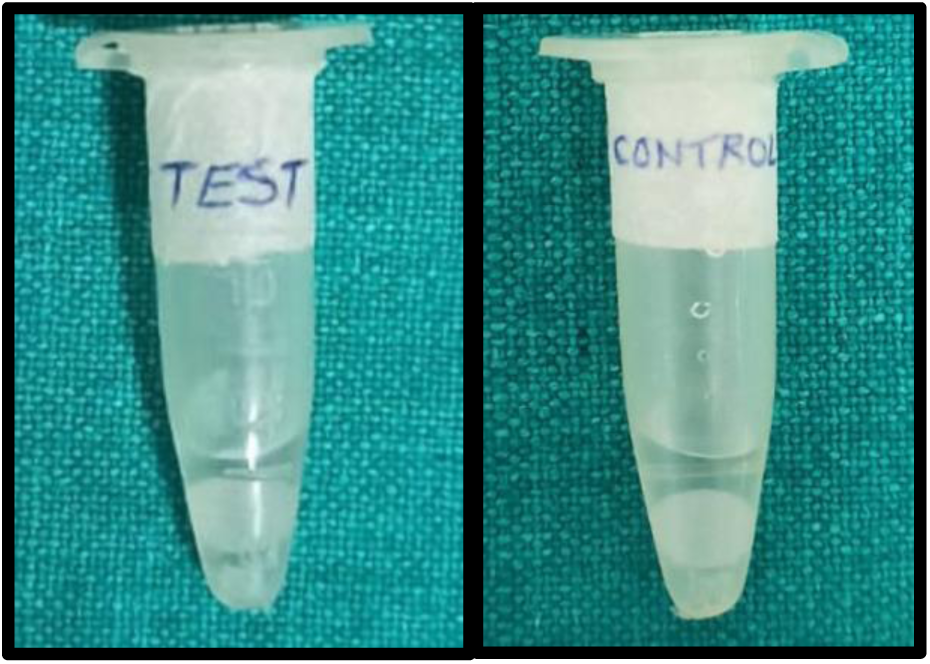
GCF Strip papers collected in Eppendorf tubes

**Fig no.10.**
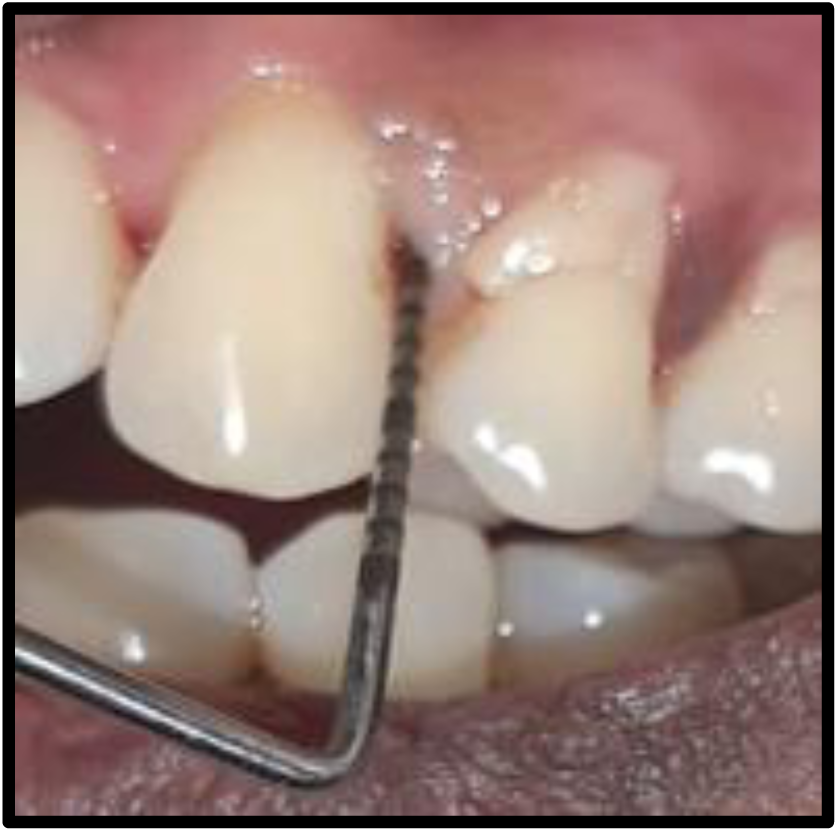
Preoperative probing depth at test site

**Fig no.11.**
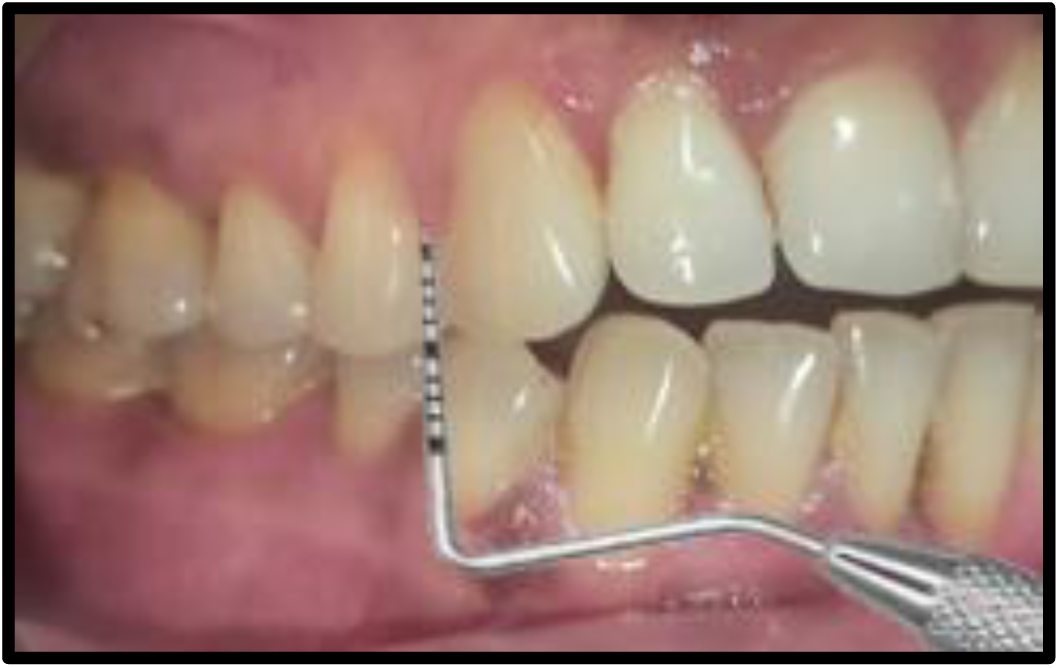
Probing depth at control site after 3 months

**Fig no.12.**
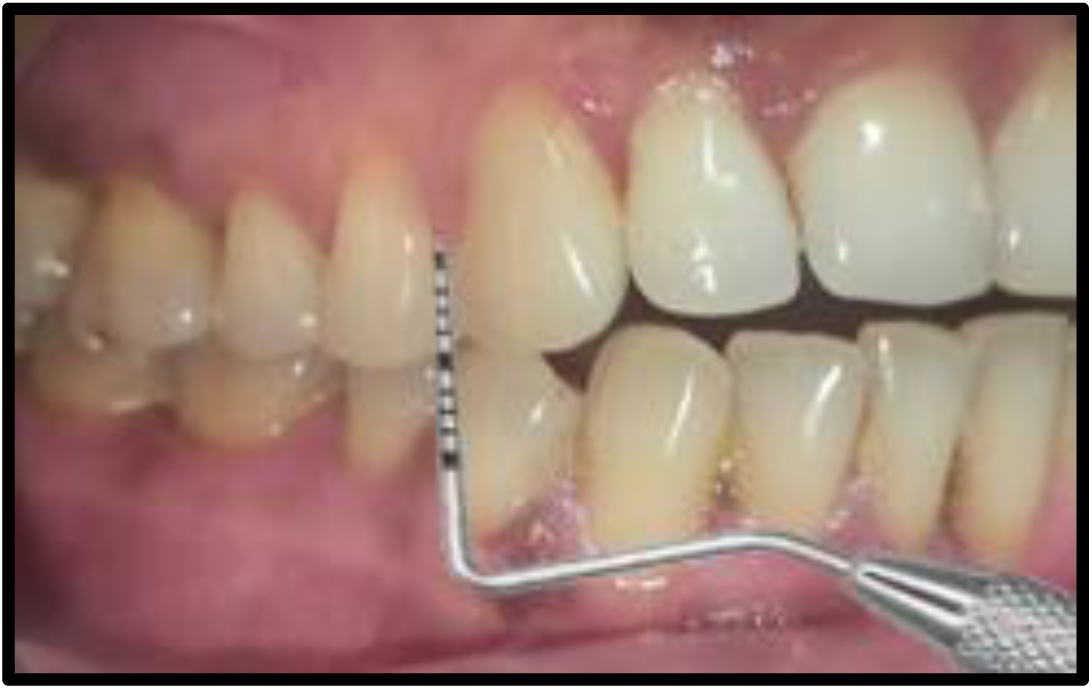
Probing depth at control site after 6 months

**Fig no.13.**
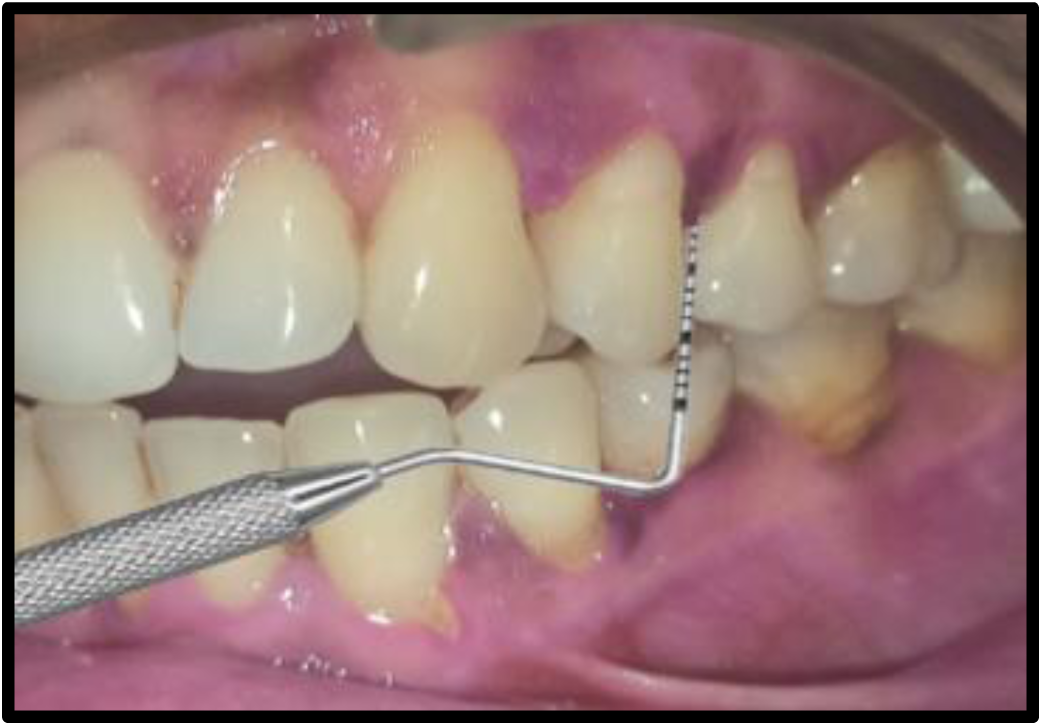
Probing depth at test site after 3 months

**Fig no.14.**
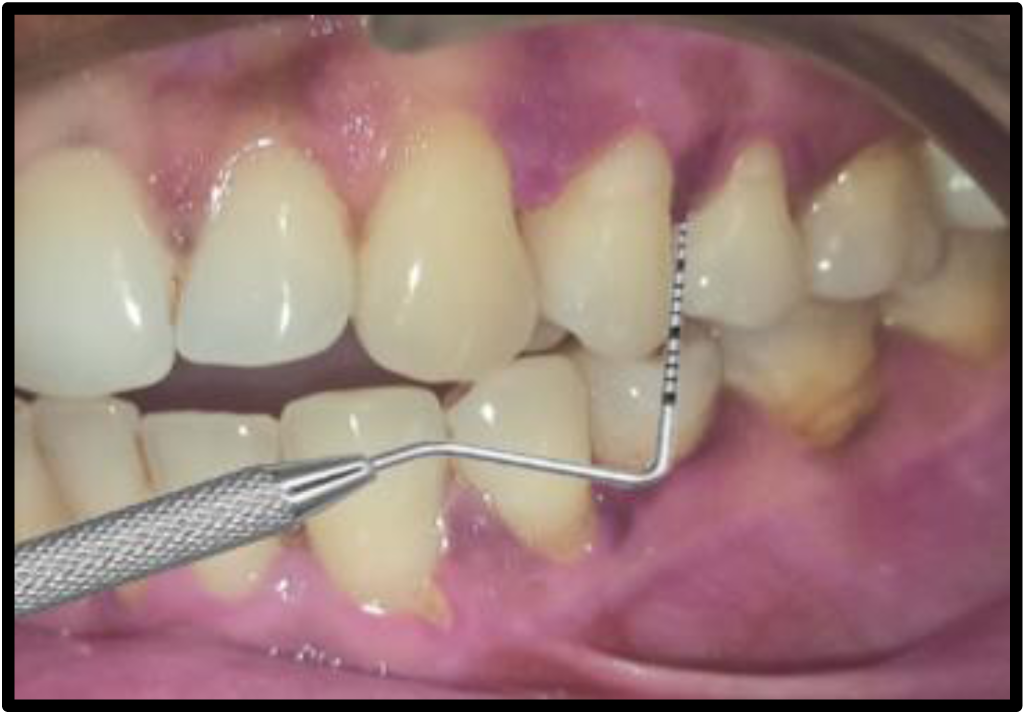
Probing depth at test site after 6 months

## Observations & Results

This split mouth randomized control trial was carried out to determine the efficacy of Stevia gel on levels of GCF glucose and ghrelin in diabetic patients with periodontitis. Nineteen subjects with established diabetes and periodontitis were recruited for this study. GCF samples of all subjects were evaluated for levels of glucose and ghrelin before and after application of Stevia gel in gingival crevice using ELISA.

### INTRAGROUP COMPARISONS

#### Probing Pocket Depth

The mean probing pocket depths (in mm) in test group were 6.52 (0.51),4.15 (0.51), 2.57 (0.50) and in the control group were 6.31 (0.48),5.21 (0.42),5.21 (0.42) at baseline, 3 and 6 months respectively (Table 1). The intra group reduction in probing pocket depth, when compared from baseline to 3 and 6 months was statistically highly significant in both the treatment groups (p˂0.001). (Table 1)

#### Clinical Attachment level

The mean clinical attachment level (in mm) in test group were 4.47 (0.51),2.57 (0.50),1.10 (0.31) and control group were 4.21 (0.42), 3.15 (0.37),3.15 (0.37) at baseline, 3 and 6 months respectively. The intra group gain in clinical attachment level, when compared from baseline to 3 and 6 months was statistically highly significant in both the treatment groups. (p<0.001). (Table 2)

#### Plaque index

The mean plaque index (in mm) in test group were 3.89 (0.31), 1.42(0.50), 1.42 (0.50), 0.31 (0.47) and control group were 3.84 (0.37), 2.57 (0.50), 2.26 (0.56) at baseline, 3 and 6 months respectively. The intra group decrease in plaque index, when compared from baseline to 3 and 6 months was statistically highly significant in both the treatment groups. (p<0.001). (Table 3)

#### Bleeding on probing

The mean bleeding on probing (in mm) in test group were 2.84 (1.12), 1.36 (1.16), 0.31 (1.37) and control group were 2.78 (0.41), 1.78 (0.41), 1.50 (0.51) at baseline, 3 and 6 months respectively. The intra group decrease in bleeding on probing, when compared from baseline to 3 and 6 months was statistically highly significant in both the treatment groups. (p<0.001). (Table 4)

#### GCF Glucose levels

The mean GCF Glucose in test group were 117.26 (8.96), 105.89 (11.1), 87.63 (5.59) and control group were 117.05 (10.12), 102.36 (11.29), 89.00 (5.95) at baseline, 3 and 6 months respectively. The intra group decrease in GCF glucose, when compared from baseline to 3 and 6 months was statistically significant in both the treatment groups. (p<0.001). (Table 5)

#### GCF Ghrelin levels

The mean GCF Glucose in test group were 102.53 (3.92), 111.66 (4.10), 118.03 (6.79) and control group were 102.23 (4.53), 113.08 (4.98), 119.05 (6.21) at baseline, 3 and 6 months respectively. The intra group increase in GCF Ghrelin, when compared from baseline to 3 and 6 months was statistically significant in both the treatment groups. (p<0.001). (Table 6)

### INTERGROUP COMPARISONS

#### Probing depth

At baseline no significant difference was observed between the two groups. At 3 months, there was significant difference in probing depth between both the groups. At 6 months however, the difference was highly significant (p˂0.001) (Table 7) (Graph 1) with stevia gel showing better probing depth reduction than placebo.

#### Clinical attachment level

At baseline no significant difference was observed between the two groups. At 3 months, there was significant difference in clinical attachment level between the groups. At 6 months however, the difference was highly significant (p˂0.001) (Table 8) (Graph 2) with stevia gel showing gain in clinical attachment level than placebo.

#### Plaque Index

At baseline no significant difference was observed between the two groups. At 3 months, there was significant difference in plaque index between the groups. At 6 months however, the difference was highly significant (p˂0.001) (Table 9) (Graph 3) with stevia gel showing better plaque index reduction than placebo.

#### Bleeding on probing

At baseline no significant difference was observed between the two groups. At 3 months, there was significant difference in bleeding on probing between the groups. At 6 months however, the difference was highly significant (p˂0.001) (Table 10) (Graph 4) with stevia gel showing better bleeding on probing reduction than placebo.

#### GCF Glucose & Ghrelin levels

No significant difference was observed between two groups from baseline to 3 months. A significant increase in GCF ghrelin levels & decrease in glucose levels was found in test group from 3 months to 6 months. (Table 11, 12) (Graph 5, 6)

### INTRA GROUP COMPARISION

**Table 1:** Intragroup comparison of mean values of probing pocket depths at baseline, 3 months and 6 months.

| PROBING DEPTH |  |  |  |  |  |
| --- | --- | --- | --- | --- | --- |
| Groups | Mean (standard deviation) |  |  | Chi square value | p-value |
|  | Base line | 3 months | 6 months |  |  |
| Test group | 6.52(0.51) | 4.15(0.51) | 2.57(0.50) | 38.00 | 0.000 HS |
| Control group | 6.31(0.48) | 5.21(0.42) | 5.21(0.42) | 35.00 | 0.000 HS |

Statistical test applied: Friedman test; HS – Highly significant at $p < 0.01$

**Table 2:** Intragroup comparison of mean values of Clinical attachment levels at baseline, 3 months and 6 months.

| **CLINICAL ATTACHMENT LEVEL** |  |  |  |  |  |
| --- | --- | --- | --- | --- | --- |
| Groups | Mean (standard deviation) |  |  | Chi square value | p-value |
|  | Base line | 3 months | 6 months |  |  |
| Test group | 4.47(0.51) | 2.57(0.50) | 1.10(0.31) | 38.00 | 0.000 HS |
| Control group | 4.21(0.42) | 3.15(0.37) | 3.15(0.37) | 35.00 | 0.000 HS |

Statistical test applied: Friedman test; HS – Highly significant at $p < 0.01$

**Table 3:** Intragroup comparison of mean values of Plaque Index at baseline, 3 months and 6 months.

| **PLAQUE INDEX** |  |  |  |  |  |
| --- | --- | --- | --- | --- | --- |
| Groups | Mean (standard deviation) |  |  | Chi square value | p-value |
|  | Base line | 3 months | 6 months |  |  |
| Test group | 3.89(0.31) | 1.42(0.50) | 0.31(0.47) | 38.00 | 0.000 HS |
| Control group | 3.84(0.37) | 2.57(0.50) | 2.26(0.56) | 35.00 | 0.000 HS |

Statistical test applied: Friedman test; HS – Highly significant at $p < 0.01$

**Table 4:** Intragroup comparison of mean values of Bleeding on probing at baseline, 3 months and 6 months.

| **BLEEDING ON PROBING** |  |  |  |  |  |
| --- | --- | --- | --- | --- | --- |
| Groups | Mean (standard deviation) |  |  | Chi square value | p-value |
|  | Base line | 3 months | 6 months |  |  |
| Test group | 2.84(1.12) | 1.36(1.16) | 0.31(1.37) | 37.52 | 0.000 HS |
| Control group | 2.78(0.41) | 1.78(0.41) | 1.52(0.51) | 34.65 | 0.000 HS |

Statistical test applied: Friedman test; HS – Highly significant at $p < 0.01$

**Table 5:**
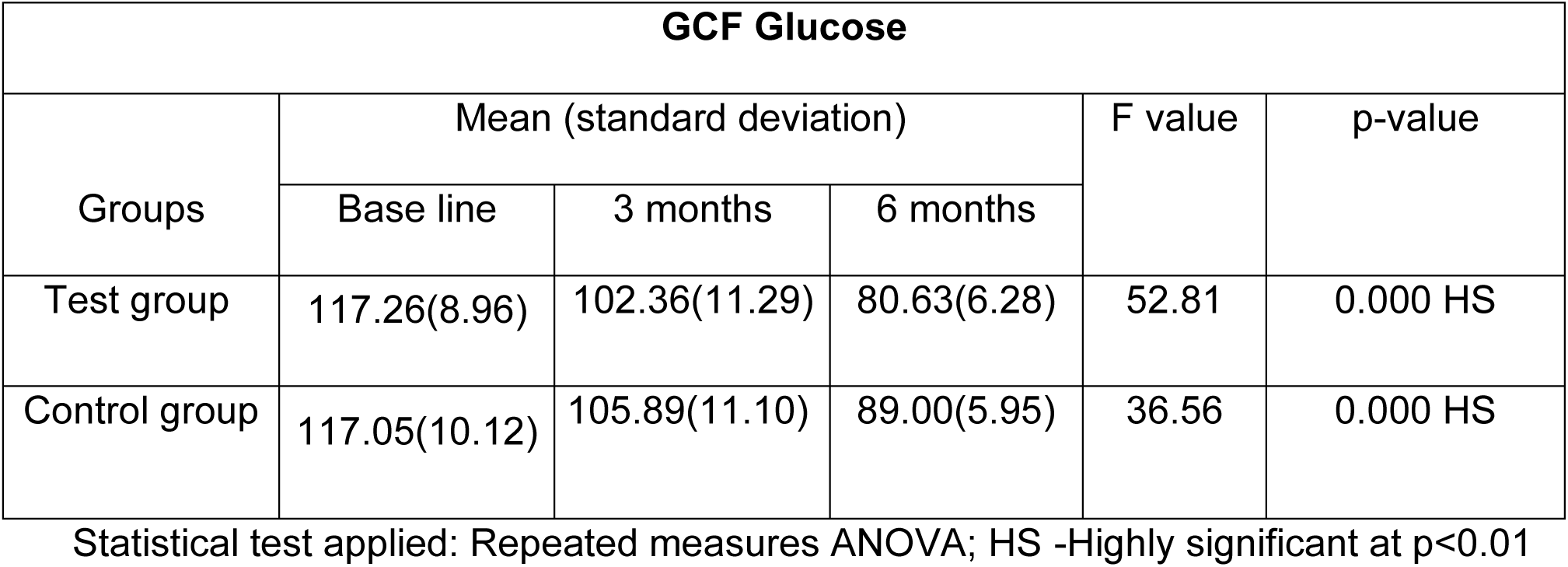
Intragroup comparison of mean values of GCF Glucose at baseline, 3 months and 6 months.

| **GCF Glucose** |  |  |  |  |  |
| --- | --- | --- | --- | --- | --- |
| Groups | Mean (standard deviation) |  |  | F value | p-value |
|  | Base line | 3 months | 6 months |  |  |
| Test group | 117.26(8.96) | 102.36(11.29) | 80.63(6.28) | 52.81 | 0.000 HS |
| Control group | 117.05(10.12) | 105.89(11.10) | 89.00(5.95) | 36.56 | 0.000 HS |

Statistical test applied: Repeated measures ANOVA; HS -Highly significant at $p < 0.01$

**Table 6:** Intragroup comparison of mean values of GCF Ghrelin at baseline, 3 months and 6 months.

| **GCF Ghrelin** |  |  |  |  |  |
| --- | --- | --- | --- | --- | --- |
| Groups | Mean (standard deviation) |  |  | F value | p-value |
|  | Base line | 3 months | 6 months |  |  |
| Test group | 102.53(3.92) | 113.08(4.98) | 130.03(11.02) | 38.69 | 0.000 HS |
| Control group | 102.23(4.53) | 111.66(4.10) | 119.05(6.21) | 55.84 | 0.000 HS |

Statistical test applied: Repeated measures ANOVA; HS -Highly significant at $p < 0.01$

### INTER GROUP COMPARISION

**Table 7:** Intergroup comparison of mean values of probing pocket depths at baseline, 3 months and 6 months.

|  | Group | N | Mean | Std. Deviation | Z value | p-value |
| --- | --- | --- | --- | --- | --- | --- |
| Baseline | Control | 19 | 6.3158 | .47757 | -1.29 | 0.27 |
|  | Test | 19 | 6.5263 | .51299 |  |  |
| 3 months | Control | 19 | 5.2105 | .41885 | -4.83 | 0.000 HS |
|  | Test | 19 | 4.1579 | .50146 |  |  |
| 6 months | Control | 19 | 5.2105 | .41885 | -5.54 | 0.000 HS |
|  | Test | 19 | 2.5789 | .50726 |  |  |

Statistical test applied: Mann Whitney U test; HS – Highly significant at $p < 0.01$

**Table 8:** Intergroup comparison of mean values of Clinical attachment levels at baseline, 3 months and 6 months.

|  | Group | N | Mean | Std.<br>Deviation | Z value | p-value |
| --- | --- | --- | --- | --- | --- | --- |
| Baseline | Control | 19 | 4.2105 | .41885 | -1.68 | 0.17 |
|  | Test | 19 | 4.4737 | .51299 |  |  |
| 3 months | Control | 19 | 3.1579 | .37463 | -3.3 | 0.006 HS |
|  | Test | 19 | 2.5789 | .50726 |  |  |
| 6 months | Control | 19 | 3.1579 | .37463 | -5.7 | 0.000 HS |
|  | Test | 19 | 1.1053 | .31530 |  |  |

Statistical test applied: Mann Whitney U test; HS – Highly significant at $p < 0.01$

**Table 9:** Intergroup comparison of mean values of Plaque Index at baseline, 3 months and 6 months.

|  | Group | N | Mean | Std.<br>Deviation | Z value | p-value |
| --- | --- | --- | --- | --- | --- | --- |
| Baseline | Control | 19 | 3.8421 | .37463 | -0.47 | 0.79 |
|  | Test | 19 | 3.8947 | .31530 |  |  |
| 3 months | Control | 19 | 2.5789 | .50726 | -4.6 | 0.000 HS |
|  | Test | 19 | 1.4211 | .50726 |  |  |
| 6 months | Control | 19 | 2.2632 | .56195 | -5.4 | 0.000 HS |
|  | Test | 19 | .3158 | .47757 |  |  |

Statistical test applied: Mann Whitney U test; HS – Highly significant at $p < 0.01$

**Table 10:** Intergroup comparison of mean values of Bleeding on probing at baseline, 3 months and 6 months.

|  | Group | N | Mean | Std. Deviation | Z value | p-value |
| --- | --- | --- | --- | --- | --- | --- |
| Baseline | Control | 19 | 2.7895 | .41885 | -0.7 | 0.54 |
|  | Test | 19 | 2.8421 | 1.11869 |  |  |
| 3 months | Control | 19 | 1.7895 | .41885 | -3.5 | 0.000 HS |
|  | Test | 19 | 1.3684 | 1.16479 |  |  |
| 6 months | Control | 19 | 1.5263 | .51299 | -5.076 | 0.000 HS |
|  | Test | 19 | .3158 | 1.37649 |  |  |

Statistical test applied: Mann Whitney U test; HS – Highly significant at $p < 0.01$

**Table 11:** Intergroup comparison of mean values of GCF Glucose at baseline, 3 months and 6 months.

|  | Group | N | Mean | Std. Deviation | Std. Error Mean | t-value | p-value |
| --- | --- | --- | --- | --- | --- | --- | --- |
| Baseline | Control | 19 | 117.0526 | 10.12957 | 2.32388 | -0.06 | 0.94 |
|  | Test | 19 | 117.2632 | 8.96191 | 2.05600 |  |  |
| 3 months | Control | 19 | 105.8947 | 11.10002 | 2.54652 | -0.97 | 0.33 |
|  | Test | 19 | 102.3684 | 11.29017 | 2.59014 |  |  |
| 6 months | Control | 19 | 89.0000 | 5.95352 | 1.36583 | 0.62 | 0.03 HS |
|  | Test | 19 | 80.6316 | 6.28622 | 1.48922 |  |  |

Statistical test applied: Independent t test; HS – Highly significant at $p < 0.01$

**Table 12:** Intergroup comparison of mean values of GCF Ghrelin at baseline, 3 months and 6 months

|  | Group | N | Mean | Std. Deviation | Std. Error Mean | t-value | p-value |
| --- | --- | --- | --- | --- | --- | --- | --- |
| Baseline | Control | 19 | 102.2316 | 4.53922 | 1.04137 | -0.22 | 0.82 |
|  | Test | 19 | 102.5368 | 3.92516 | 0.90049 |  |  |
| 3 months | Control | 19 | 111.6684 | 4.10948 | 0.94278 | 0.95 | 0.34 |
|  | Test | 19 | 113.0895 | 4.98051 | 1.14261 |  |  |
| 6 months | Control | 19 | 119.0526 | 6.21444 | 1.42569 | 0.39 | 0.002 HS |
|  | Test | 19 | 130.0316 | 11.0282 | 9.63921 |  |  |

Statistical test applied: Independent t test; HS – Highly significant at $p < 0.01$

**Graph 1:**
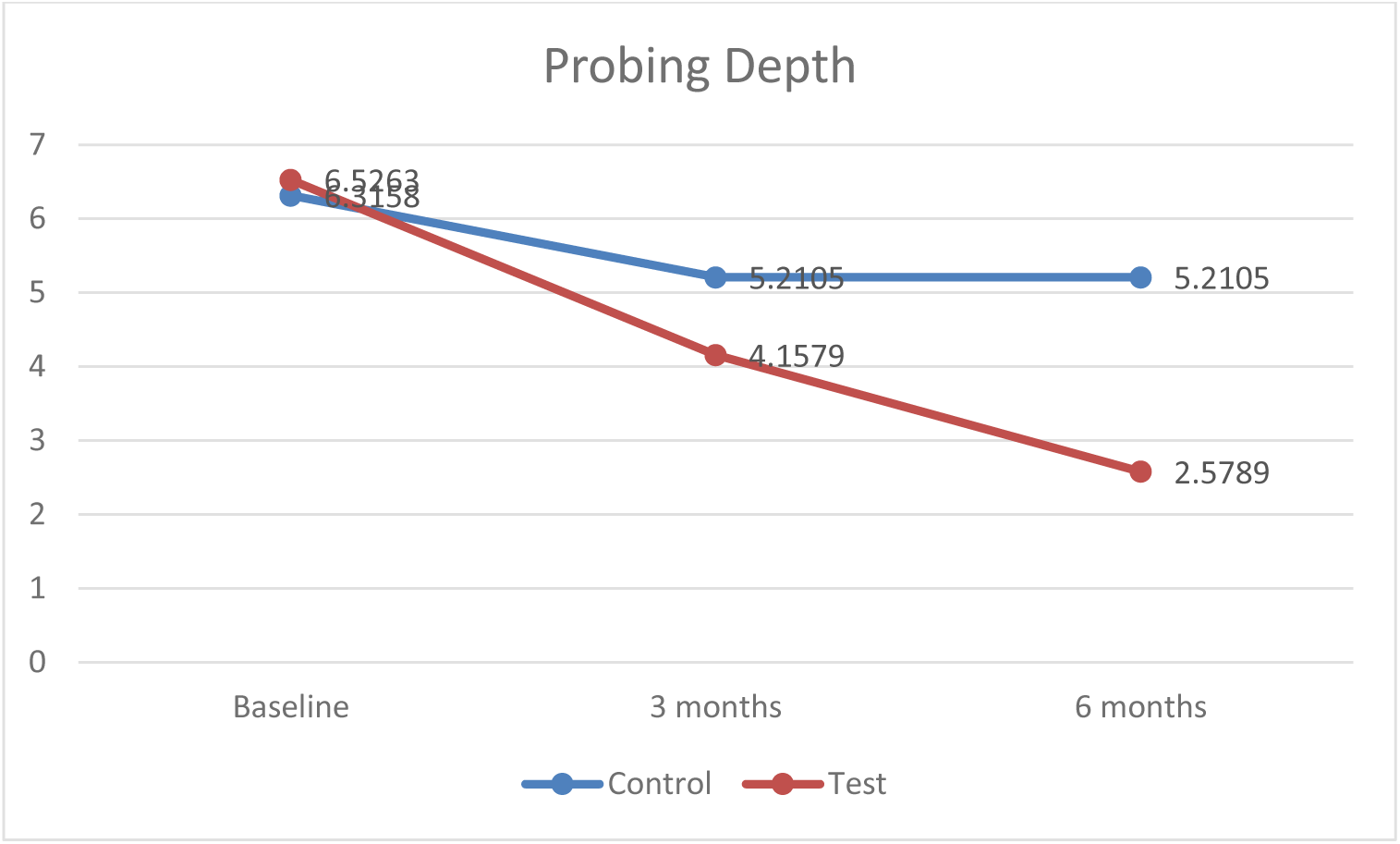
Intergroup comparison of mean values of probing pocket depths at baseline, 3 months and 6 months.

**Graph 2:**
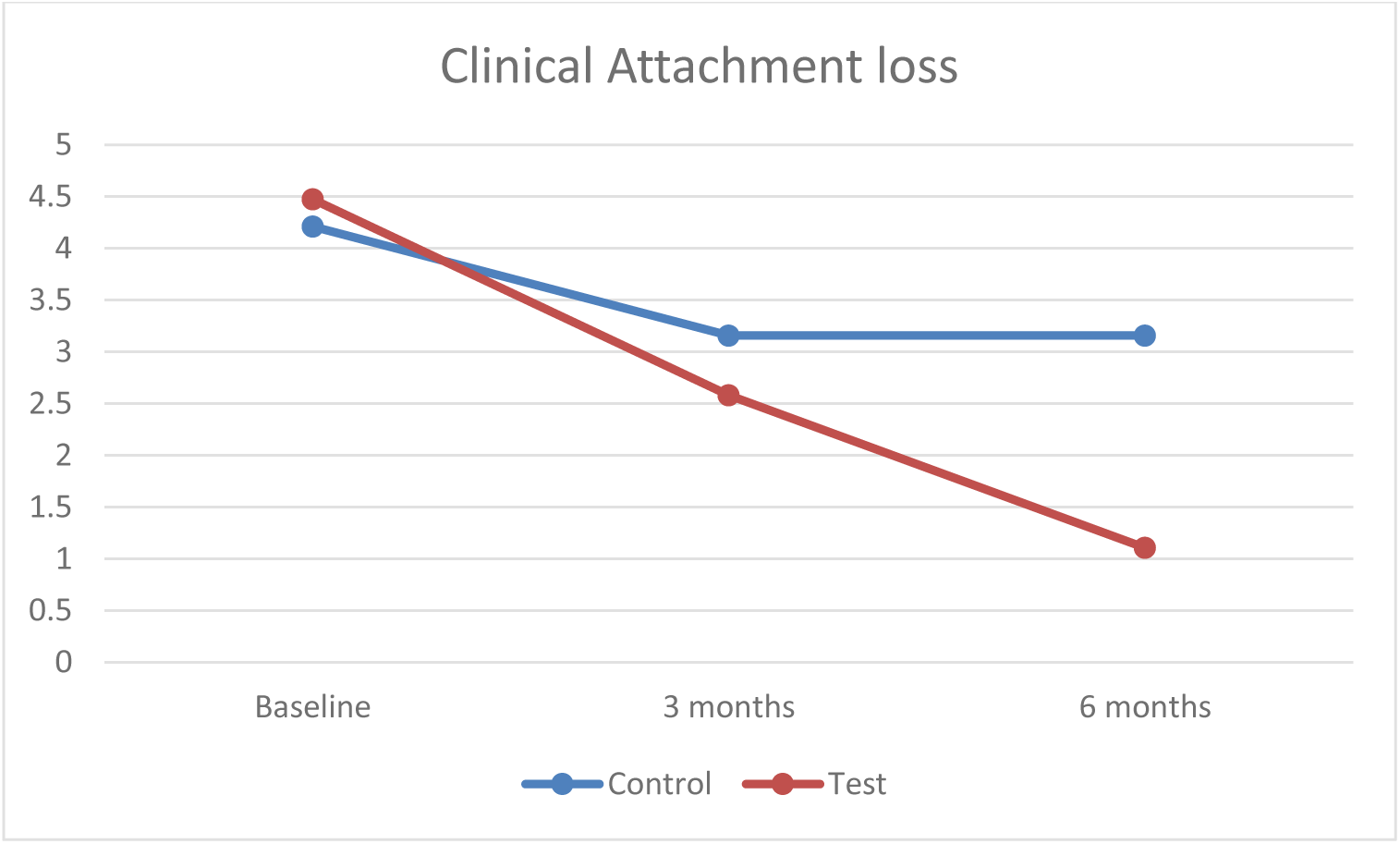
Intergroup comparison of mean values of clinical attachment levels at baseline, 3 months and 6 months.

**Graph 3:**
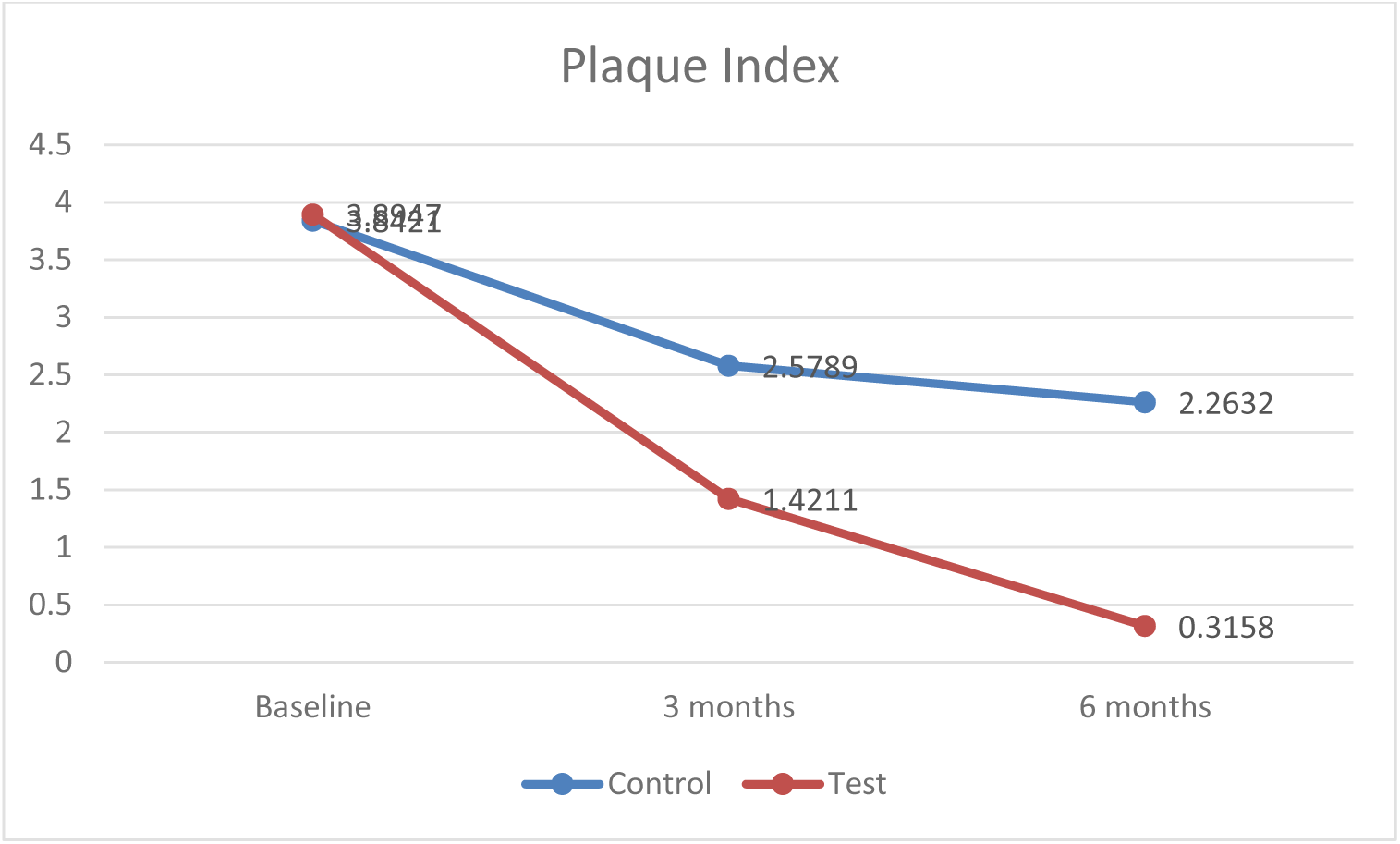
Intergroup comparison of mean values of plaque index at baseline, 3 months and 6 months.

**Graph 4:**
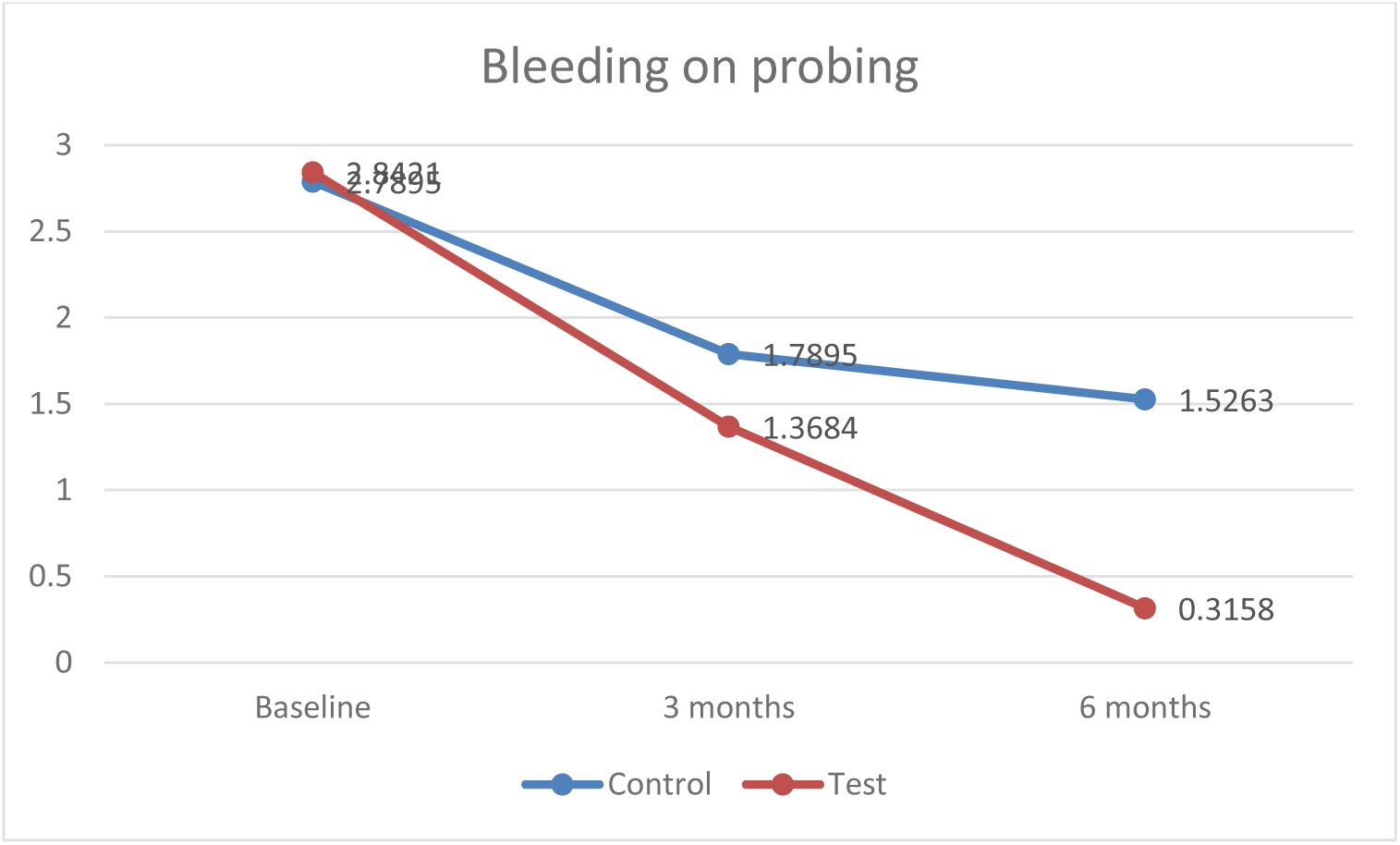
Intergroup comparison of mean values of bleeding on probing at baseline, 3 months and 6 months.

**Graph 5:**
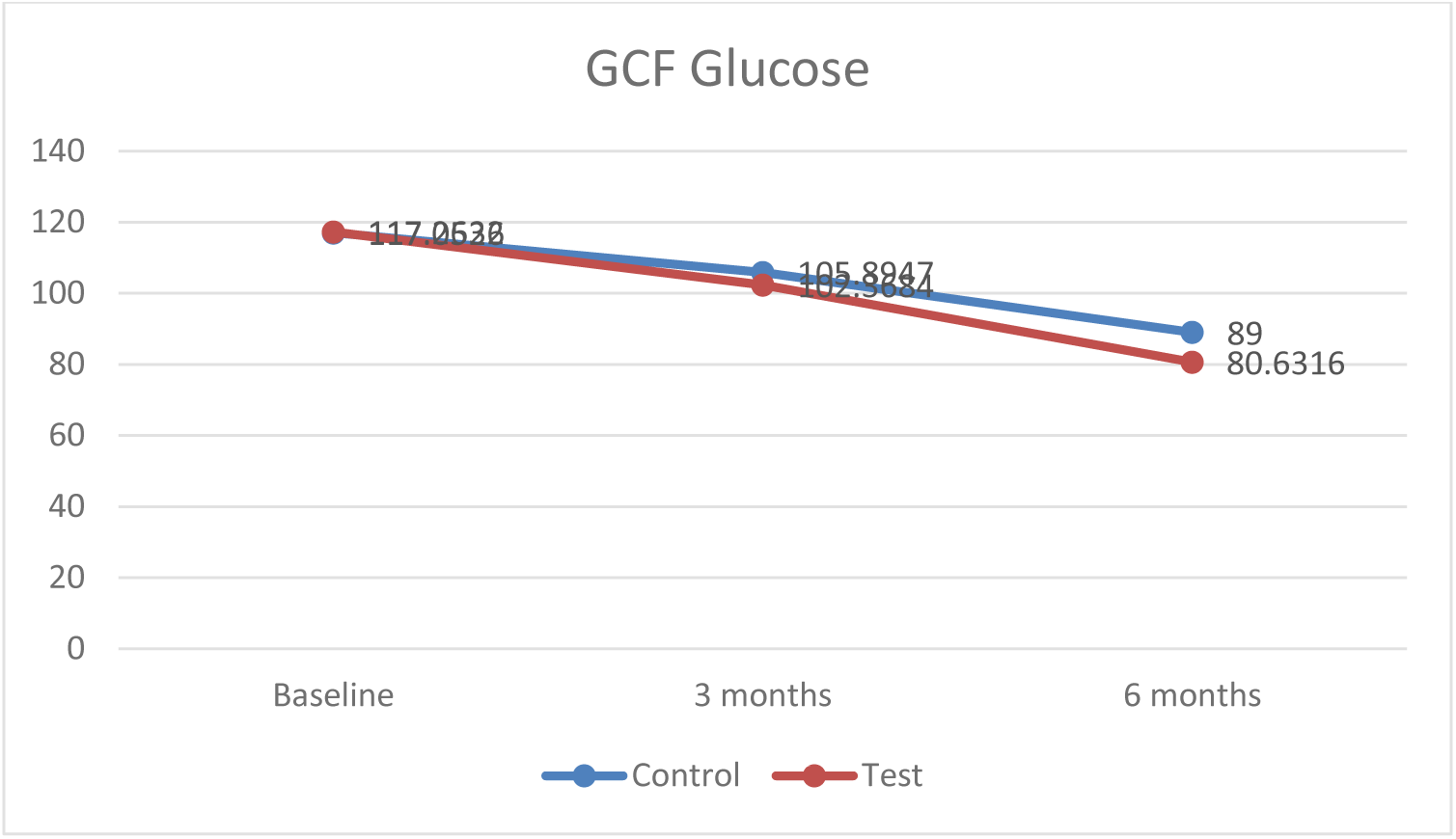
Intergroup comparison of mean values of GCF Glucose at baseline, 3 months and 6 months.

**Graph 6:**
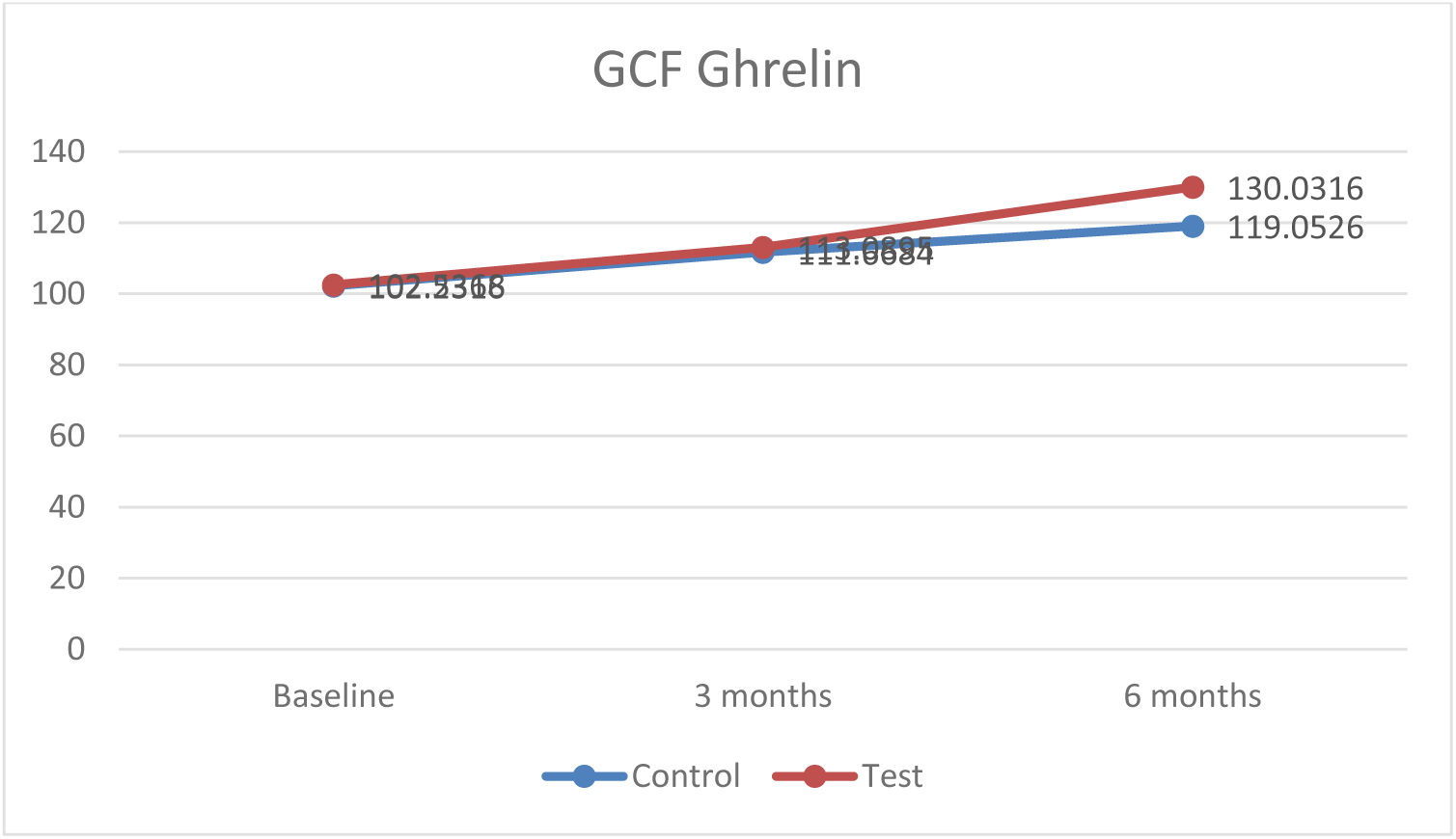
Intergroup comparison of mean values of GCF Ghrelin at baseline, 3 months and 6 months

## Discussion

Periodontitis being a common inflammatory condition of the supporting structures of the teeth and is epitomized by an exacerbated production of pro-inflammatory molecules that result in the breakdown of the extracellular matrix and alveolar bone destruction.^2^ While the associations between periodontal diseases and various systemic disorders have been validated through longitudinal studies, the bond with diabetes mellitus is deep-rooted in medical evidence. Multiple controlled trials and systemic reviews have established diabetes mellitus to be the strongest risk factor for inflammatory periodontal disease.^5^ The presence of untreated periodontal disease in turn, is known to negatively influence the hyperglycemic state. The dysregulated immune functions and exaggerated inflammatory reactions account for the elevated levels of inflammation-promoting biomolecules in various body fluids, in patients with diabetes mellitus. There is thus an overwhelming and incontrovertible cache of evidence demonstrating a bidirectional association among these two common inflammatory conditions-diabetes mellitus and periodontitis, where in the common denominator between the two diseases seems to be a hyperinflammatory trait.^4,6^

Periodontitis is a chronic disease that frequently undergoes short periods of activity that is followed by longer periods of quiescence. It would therefore be sensible to identify corresponding biomarkers that estimate the extent of activity of the inflammatory process to aid in the detection of periodontitis and to evolve appropriate treatment strategies based on the insight of the current state of disease activity. Gingival crevicular fluid (GCF) is a transudate as well as an inflammatory exudate originating from the gingival plexus of blood vessels in the gingival connective tissue which is present subjacent to the epithelium lining the gingival sulcus. It is composed of serum and locally generated components such as tissue breakdown products, inflammatory mediators and antibodies in response to oral microorganisms present in the dental plaque. Thus, it offers great potential to reflect the response that the cells and periodontal tissues promote to attempt regaining homeostasis and also how certain Periodontol pathogens co-opt these response mechanisms to promote bacterial survival within the gingival crevice and periodontal pocket. GCF also has other favorable features such as, ease of access, requirement of minimal operator training etc, that made GCF the tissue fluid of choice to assess for biomarkers that pertain to oral inflammatory disease.^49,50^.

India was found to be fast-developing economy with a considerable number of diabetic patients. It was recorded as second highest country after China with 65.1 million diabetes cases which was estimated in 2013. This can be expected to increase up to 109.0 million cases till 2035.^51^

A native plant *Stevia rebaudiana* (SR) from South America was found with medicinal values since long time. It has two main glycosides that are stevioside (110-270 times sweeter than sugar) and rebaudioside A (180-400 times sweeter than sugar), the last one with higher commercial valuable, because it shows a nice flavour profile, unlike artificial sweeteners such as saccharin, aspartame, acesulfame-K, and cyclamate that have metallic taste^56^ or the same stevioside that has a subtle bitter flavour.^53,54^ The only difference between these glycosides lies in the presence of a glucose and its fraction of weight in the tissues of the plant.^52,53,54^ It was used as a natural sweetener with zero caloric value in popular foods and beverages. Furthermore, it was used for various therapeutic purposes such as an antacid, antimicrobial, antioxidant, and antidiuretic.^55^

Steviol glycosides was also found not to induce a glycaemic response when it was ingested, which had made them attractive as zero or low-calories natural sweeteners to diabetics and other people with carbohydrate-controlled diets. Various studies results had demonstrated that the treatment with Stevia increased the tolerance to glucose and decreased concentrations of plasmatic glucose.^59^ Evidence by **Tarka et al**^57^ suggested that stevioside improves secretion and sensitivity to insulin. Besides, it was found that it generates the concomitant suppression of glucagon’s secretion and declines the renal tubular reabsorption of glucose.^58^

Diabetes was characterized by hyperglycaemia, which affects the cells indirectly, through the production of AGEs and directly through the stimulation of different intracellular pathways that resulted in several cellular changes. Hyperglycaemia (increased glucose) also contributed to the pathogenesis of periodontitis in diabetes.^60,61^ A peptide hormone Ghrelin was principally secreted by the stomach and was found in lesser amount in the other tissues of the body including salivary glands and teeth. Various studies have shown that ghrelin plays a major role in the gastrointestinal tract, stimulating gastric contractility and acid secretion and it is responsible for the metabolic response to starvation by modulating insulin secretion, glucose metabolism, and amino acid uptake. Furthermore, Ghrelin showed anti-inflammatory activity by decreasing pro-inflammatory cytokine production such as interleukin IL-1b and tumor necrosis factor (TNF-α) by down regulating the activity of lipopolysaccharide (LPS). ^62^

Therefore, assessing these biomarkers together by application of Stevia gel in gingival sulcus could provide a clear insight on the risk of disease progression in all the subjects as each of these markers represent various channels of the disease process.

This study was designed to evaluate the efficacy of Stevia gel for downregulation / upregulation of these markers. Test group consisted of diabetic subjects with periodontitis where Stevia gel was used after SRP. Control group consisted of diabetic subjects with periodontitis where placebo was used after SRP.

The GCF samples acquired from all the participants showed detectable range of glucose and ghrelin when evaluated using ELISA. On overall analysis, the levels of glucose were found to decrease at both test (SRP + Stevia gel) and control sites (SRP + Placebo) which was comparable to a study by **Allen et al**^63^ where periodontal disease has a devastating effect on glycemic control among type 2 diabetic patients and a significant reduction of Glycated hemoglobin (HA1c), 0.40 %, was observed after 3–4 months of periodontal therapy done **i**n patients with both type 1 and 2 DM and periodontitis.^63^

Similar results were found in a systematic review and meta-analysis by **Teshome et al**^64^, where a significant reduction of Glycated haemoglobin and Fasting plasma glucose level were found in type 2 diabetic and periodontal patients with non-surgical periodontal therapy.^64^

Another study by **Mauri-obradors et al.**^65^ stated that non-surgical periodontal treatment resulted in a better glycaemic status in type 2 diabetic patients and demonstrated the importance of oral health in their general health.

Our study results showed a significant reduction in glucose levels in control group after scaling and root planing. These results were in accordance with a study by **Munjal et al.**^66^ showed that HbA1c levels were more significantly reduced in test group, who received antibiotic coverage with non-surgical periodontal therapy (scaling and root planning) compared to the other group, who received only scaling and root planning.

In contrast to these results a study by **Mizuno et al.**^67^ stated that changes in HbA1c levels were not significant in the test group who received periodontal treatment when compared to the control group who did not receive periodontal therapy at 3 and 6 months.

A study by **Hsu et al.**^68^ stated that diabetes mellitus (HbA1c ≤ 8.5%) does not appear to significantly affect short-term clinical periodontal outcomes of non-surgical periodontal treatment.

Intergroup comparisons showed a statistically significant difference (p < 0.001) was found in glucose levels at 6 months following application of Stevia gel in test group (80.63 ± 6.28) compared to the control group (89.00 ± 1.48), where only SRP was performed (Table 11 & Graph 5). These results were comparable to a study by **Kujur et al.**^69^ stating that Stevia significantly reduced mean serum glucose levels in diabetic rats over a 1-month period when compared to controls (treated with glyburide).^77^ Similar findings were also observed in a study by **Jeppenson et al.**^74^ where experimental group, who received intervention with Stevia leaves was found to be statistically significant (p < 0.001) when compared to the control group who did not receive Stevia intervention.

An animal study by **Chang et al**^71^ found that Stevia increases insulin sensitivity in rodent models and to have beneficial effects on blood glucose.^81^ A human study by **Gregersen et al.**^32^ showed a significant reduction in mean post prandial blood glucose levels after administration of stevioside when compared to controls.

An experiment by **Saravanan and Ramachandran et al.**^70^ examined the effects of rebaudioside A on blood glucose and insulin levels, lipid peroxidation, antioxidative activity, and lipid profile on healthy and diabetic Wistar rats and was found that the treatment with rebaudioside A improved blood glucose and insulin levels in diabetic rats. Stevia was also found to cause hypoglycaemia in patients with diabetes. The possible glucose lowering action may be due to the direct impact of stevioside on pancreatic beta cells to secrete more insulin and to improve their function in glucotoxicity.^74^ In addition to its properties such as antihyperglycemic and antioxidative, its glycosides were detected in several tissues such as kidney, liver and pancreas.^73^ Scientists have also reported that the glycosides in Stevia have insulinotropic effects and may serve a potential role in the treatment of type 2 diabetes mellitus.^75^

Present study showed an inverse relation between glucose and ghrelin levels. The data was supported by a series of other studies^77,78,79^ which demonstrated increased plasma levels of glucose by decreasing insulin levels following administration of ghrelin. Notably, a link between ghrelin and insulin is also suggested by the fact that both hormones exhibit a reciprocal correlation with insulin levels being high when ghrelin levels are low and vice versa.^78^ Also, epidemiological studies support the inverse relationship between ghrelin and indexes of impaired glucose tolerance and insulin resistance.^79^

A study by **Poher et al.**^80^ stated that ghrelin morphs into a vanguard for blood glucose control during extreme settings of metabolism and nutrition. In addition to that ghrelin’s glucoregulatory actions have evolved as a system to support blood glucose during extreme negative energy balance such as starvation, during which its plasma levels naturally increase. Alternatively, ghrelin’s actions can be diabetogenic during diet-induced obesity and some monogenic forms of diabetes.^80^

Similarly other study by **Banks et al**.^81^ stated that Ghrelin has a role in maintaining blood glucose levels in starvation because it plays a paracrine role in the pancreas in the control of insulin release. Ghrelin was found to counteract the consequences of diabetes induced by oxidative stress and play an antiapoptotic and proliferative role in the pancreatic b-cell.

An experimental study **Poykko et al.**^82^ demonstrated that glucose administration or food intake have shown to decrease plasma ghrelin concentrations. In addition to that ghrelin was found to show both stimulatory and inhibitory effects on insulin secretion. In human subjects, insulin infusion has shown a decrease in ghrelin concentrations, whereas parenteral administration of insulin had no effect on ghrelin concentrations.^82^

Our study showed an increased GCF ghrelin levels at both test and control sites but more significant increase was found in test group from baseline **(102.53 ± 3.92)** to 6 months **(130.03 ± 11.02)** when compared to control group. These findings were comparable to a series of studies by **Mohamed et al**^14^ and **Yilmaz et al.**^40^ where ghrelin levels were significantly increased in patients with chronic periodontitis and T2DM than in individuals without T2DM.

An animal model by **Stengel et al.**^87^ showed that bacterial lipopolysaccharides significantly reduce the levels of desacyl ghrelin and the expression of the ghrelin acylating enzyme, ghrelin O-acyltransferase, resulting in a remarkable decrease of acylated ghrelin.^100^

Various studies by **Wang et al,**^88^ **Huang et al,**^89^ **Au et al,**^90^ **Waseem et al.**^91,92^ demonstrated that IL-1β decreased plasma ghrelin levels, whereas the application of ghrelin inhibited the inflammatory response by decreasing levels of IL-1β and tumour necrosis factor-α and augmented anti-inflammatory effects as shown by an increased expression of anti-inflammatory M2-type macrophages and an increased release of the anti-inflammatory cytokine, IL-10. Ghrelin has the potential in controlling periodontitis because of its positive modulatory effects on the immune system.

A significant reduction (p < 0.001) in PI and CAL was observed in our study, using Stevia gel showed from baseline to 6 months (Table 8 & 9). These findings were in accordance with a study by **Tiwari et al,**^83^ where Stevia mouthwash showed a significant reduction in plaque and gingival scores from the baseline to till 8^th^ day.

Similar results were also found in a study done by **Vandana et al.**^11^ showed reduction in mean plaque scores using 10% Stevia mouthwash at 3 months and 6 months as compared from the baseline. The anti-plaque effect of Stevia can be attributed to the presence of tannins and xanthine’s (theobromine and caffeine), and antigingivitis effect can be attributed to the presence of flavonoids which possess anti-inflammatory activity.^11^

In a comparative study by **De Slavutzky et al**^84^ demonstrated that formation of dental plaque was less in 100 percent of the cases with Stevia when compared to sucrose & the mean difference was of 82% less plaque for Stevia when the Silness-Löe Index was used and 40% with the ÓLeary Index. Thus, it can act as an anti-cariogenic, anti-gingivitis product and be a helper in obesity.^84^

A Study by **Lin et al.**^85^ stated that Stevia has ability to inhibit the growth of certain bacteria, which explains its use in the treatment of wounds, sores and gum diseases, besides this it also contributes to its anti-inflammatory and antiplaque effect.^85^

Intergroup comparisons showed a significant decline in BOP & PPDs with application of Stevia gel at test sites when compared to control sites from baseline to 6 months (Table 7&10). These results were in line with a study by **Bolanos et al.**^86^ where effects of Stevia extracts were observed in periodontal disease. Furthermore, it showed a significant reduction in the rate of gingival bleeding that initially ranged from 65% - 80% and after the treatment it was reduced from 12% - 10% with reduced periodontal probing depths of 4mm.^86^

Our study results demonstrated reduction in clinical parameters (BOP, PI, CAL, PPD) in both test and control group but a significant reduction was found in test group compared to control group from baseline to 6 months.

These results were comparable to a study by **Aziz et al.**^96^ which demonstrated the efficacy of SRP in effectively eliminating the etiologic agent, thereby helping in decreasing the inflammatory impact on the periodontal tissues and subsequently improving clinical parameters (BOP, PD, CAL, PI), systemic biochemical oxidative stress and inflammatory markers.

Our study findings showed a significant increase in ghrelin levels with the application of Stevia gel in test group compared to control group from baseline to 6 months.

These findings were in accordance with a study by **Akki et al.**^93^ which demonstrated that ghrelin is an endogenous antioxidant and functions as a free radical scavenger. It also inhibits apoptosis by down-regulation of Bax, preventing cytochrome c release, and inhibition of ROS formation, increasing antioxidant enzyme activities, and reducing lipid peroxidation.

A study by **Ruiz et al.**^97^ stated that the high antioxidant activity of *Stevia rebaudiana Bertoni* enhanced the potential interest in natural source of sweeteners to improve the efficacy of different products as nutraceutical and pharmacological agents. The consumption of the Stevia rebaudiana Bertoni may play a role in preventing human diseases in which free radicals are involved such as cancer, cardiovascular disease, and aging.

Other study by **Ameer et al.**^98^ stated that in addition to providing sweetness, Stevia exhibits strong antioxidant potential as a sugar substitute owing to the presence of various compounds with medicinal significance such as phenolic compounds, flavonoids, diterpene glycosides (stevioside and rebaudioside-A), condensed tannins, anthocyanins, and phenolic acids.

A single biomarker cannot be predictive of periodontitis owing to the complexity of the disease which calls for a “fusion of biomarkers” together with clinical evaluation Increased precision on using biomarker combinations have also been emphasized by **Almehmadi et al.**^94^ and **Sukriti KC et al.**^95^ in their systematic reviews.

To our best knowledge, this is the first study to compare the levels of GCF glucose and ghrelin before and after application of Stevia gel in diabetic patients with periodontitis. All studies in the literature have investigated effect of various forms of Stevia such as powder, mouth rinses and diet in periodontitis and diabetes mellitus. The nature and magnitude of the influence of Stevia gel in diabetic patients with periodontitis has not yet been understood completely where, this study might possibly fill that lacuna, at least in part.

The present study is also subjected to limitations, confounding factors such as lifestyle, environmental factors which may influence biomarkers in diabetic patients were not considered in this study. Long-term studies with multi-center trials comparing the levels of these GCF markers before and after application of Stevia gel in treatment of diabetic patients with periodontitis, with a large sample size in various geographical areas and ethnic groups should be done. Furthermore, long-term studies also needed to be carried out in the future to justify the potential diagnostic and probable prognostic value of these biomarkers or as part of treatment of diabetes and periodontal diseases.

## Conclusion & Summary

The current study evaluated the efficacy of Stevia gel on GCF levels of glucose and ghrelin in diabetic patients. Their expression was also compared and correlated among all groups and with clinical parameters. Nineteen subjects were enrolled in this study and were randomly assigned as test site for Stevia gel placement, where as other site was assigned for placebo placement as control. GCF samples were acquired and analyzed for estimation of glucose and ghrelin levels using ELISA. Following conclusions were made from this study:

- Ghrelin levels were increased in subjects after application of stevia gel in gingival crevice from base line to 6months.
- Glucose levels were decreased from baseline to 6 months in subjects after application of stevia gel in gingival crevice.
- Clinical parameters like PD, CAL, BOP, PI were significantly decreased with application of stevia gel in gingival crevice.

This study unveils the potential of Stevia gel in regulating biomarkers (glucose & ghrelin) which indicates the inflammatory burden in diabetic patients with periodontitis. It also emphasizes the colossal influence of diabetes on the upregulation of inflammatory periodontal disease progression reactions and thereby. Our results further highlight the importance of regular supportive care maintenance for diabetic patients in dental clinics. The study also strengthens the role of the less investigated yet potential Stevia gel in the regulation of periodontal inflammation in diabetic individuals.

Future long-term studies are required to validate these findings and transform the use of these glucoregulatory biomarkers into implementation in clinical forum. These biomarkers apart from being of diagnostic purpose, also appear to be promising prognostic markers and therapeutic targets for periodontal disease & diabetes which needs to be explored.

## Data Availability

All data produced in the present study are available upon reasonable request to the authors

## Annexures

### ANNEXURE I INFORMED CONSENT

I ____________________________________ the undersigned hereby give my consent for undergoing the treatment as a part of the dissertation titled; “**EFFICACY OF STEVIA REBAUDIANA BERTONI ON LEVELS OF GCF GLUCOSE AND SELECTED BIO-MARKERS IN DIABETIC PATIENTS WITH PERIODONTITIS-A SPLIT MOUTH RANDOMOIZED CONTROLLED TRIAL**” being conducted by Dr. K. NANDINI under the guidance of DR. A. AMARENDER REDDY MDS, PROFESSOR, Department of Periodontics, SVS Institute of Dental Sciences. I have been explained about the details of the procedure in my vernacular language, which I understood and hereby voluntarily, unconditionally give my consent without any fear or pressure in mentally sound and conscious state to participate in this study.

Sign __________

### ANNEXURE II EFFICACY OF STEVIA REBAUDIANA BERTONI ON LEVELS OF GCF GLUCOSE AND SELECTED BIO-MARKERS IN DIABETIC PATIENTS WITH PERIODONTITIS-A SPLIT MOUTH RANDOMOIZED CONTROLLED TRIAL

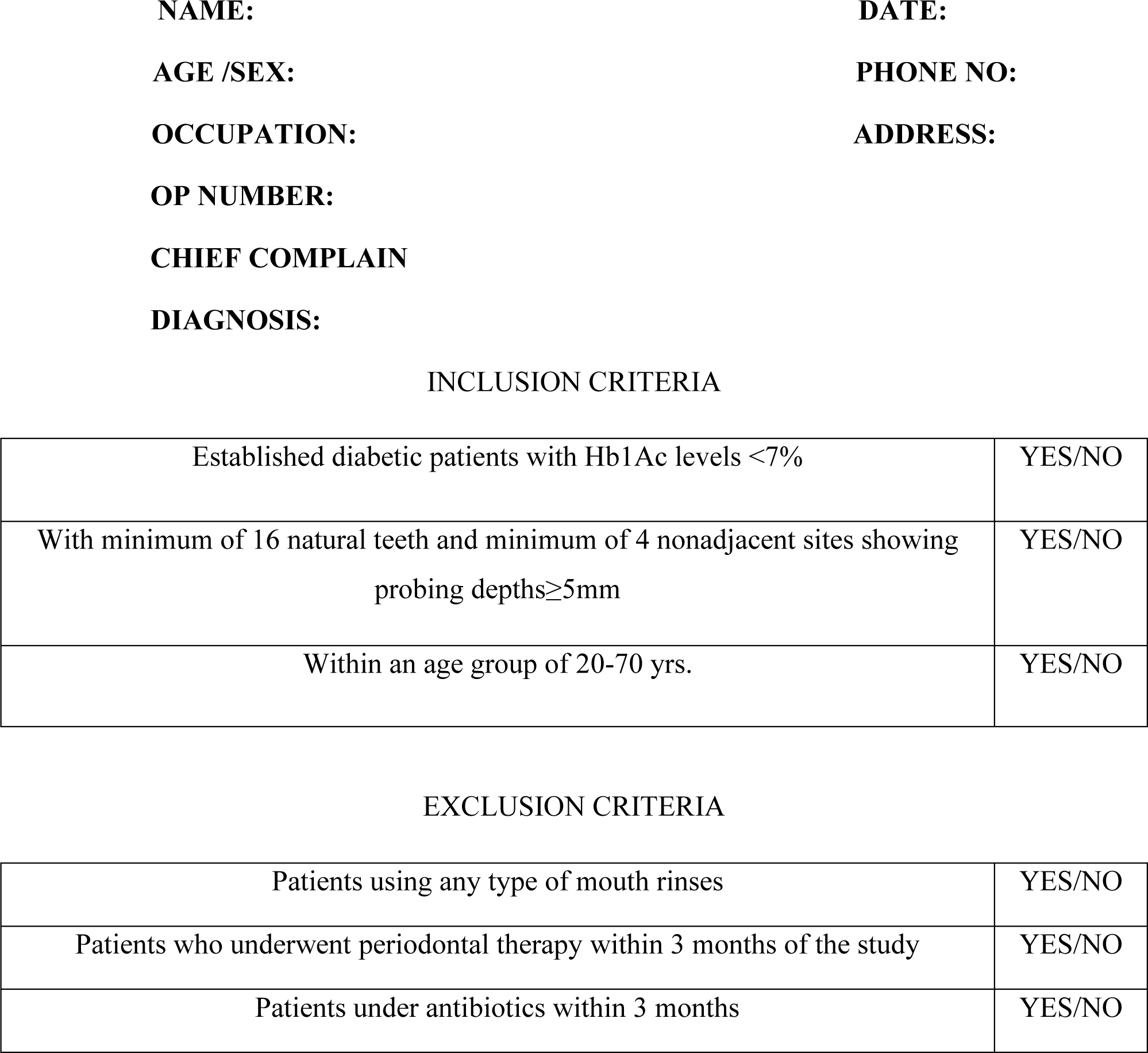

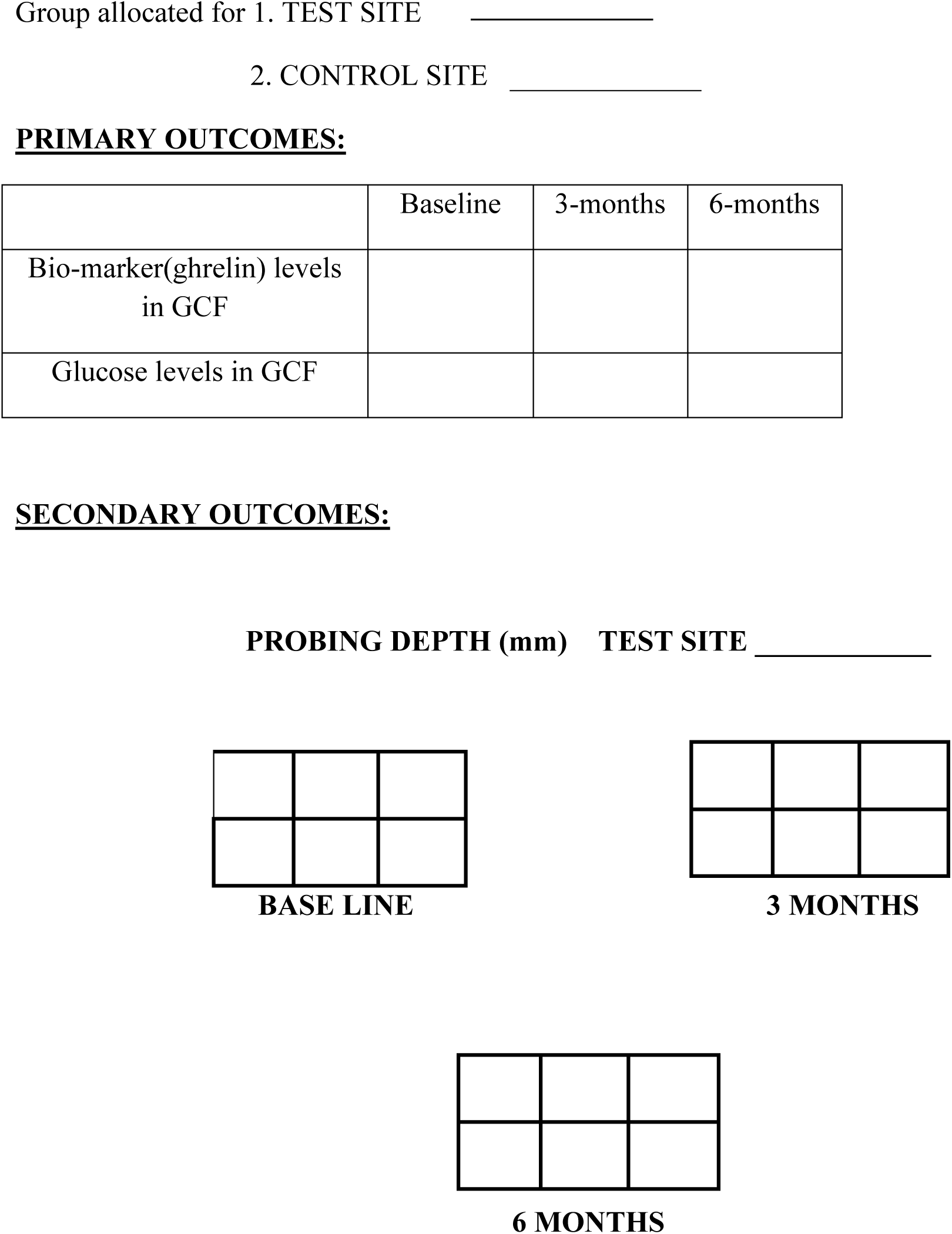

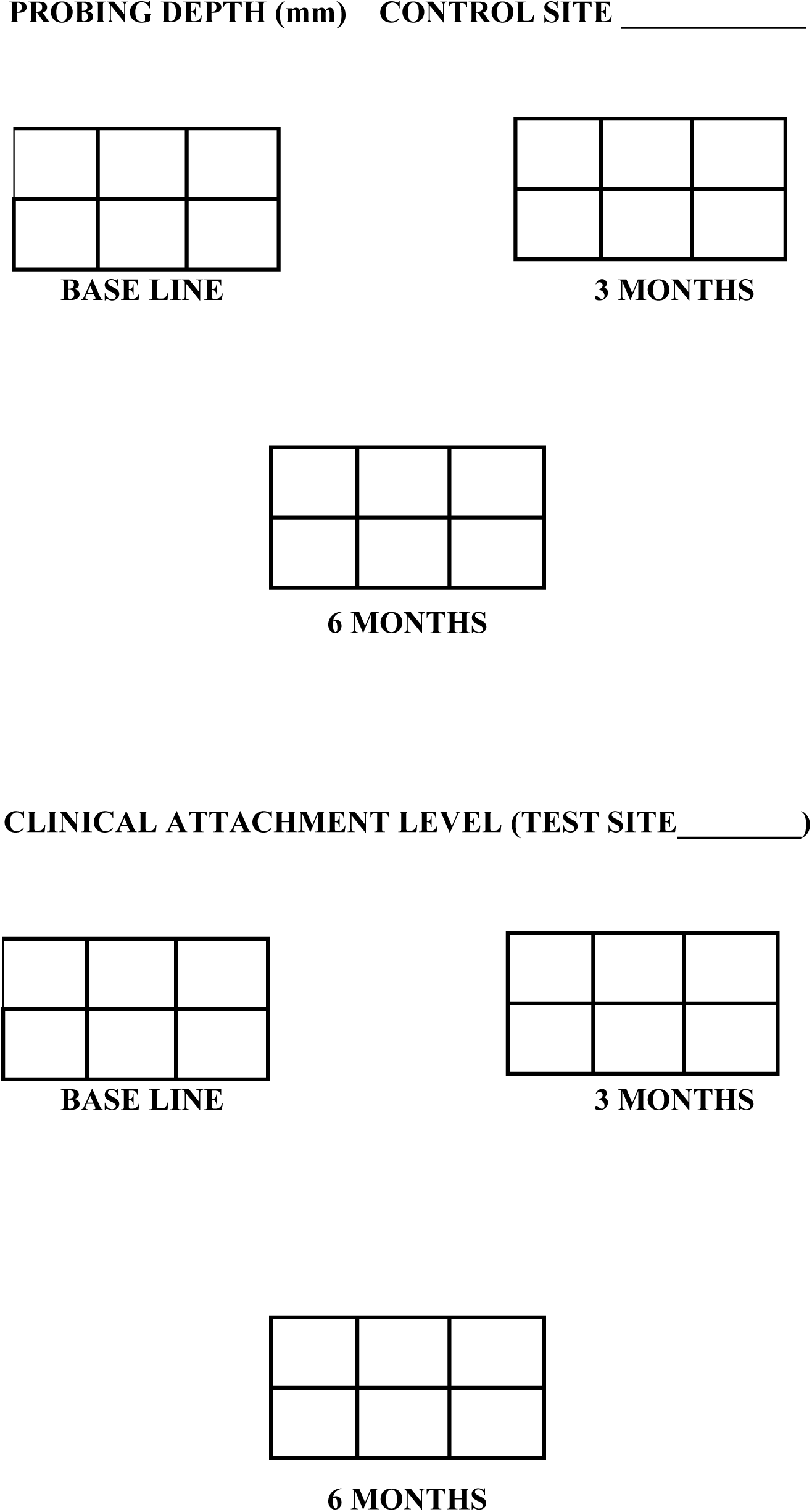

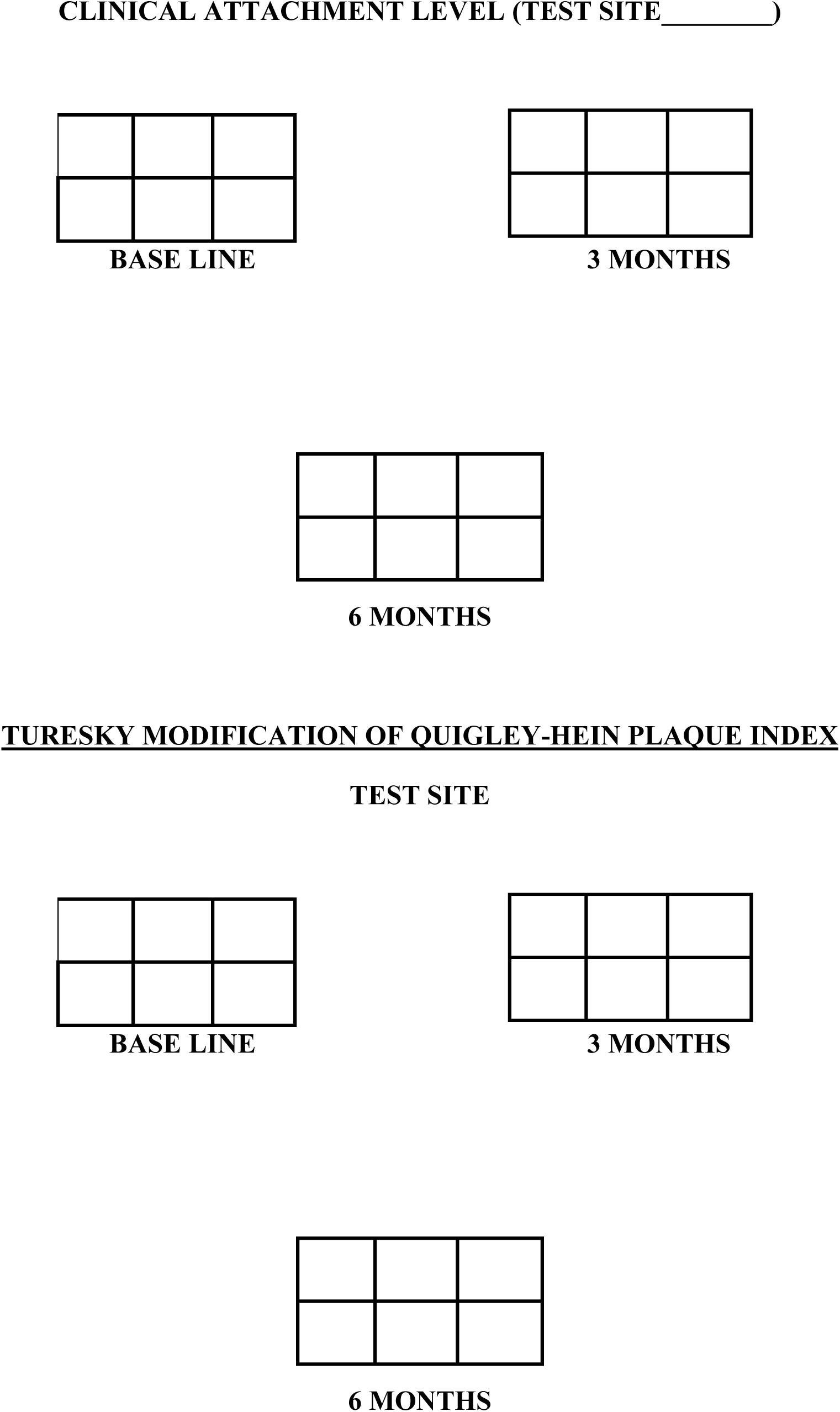

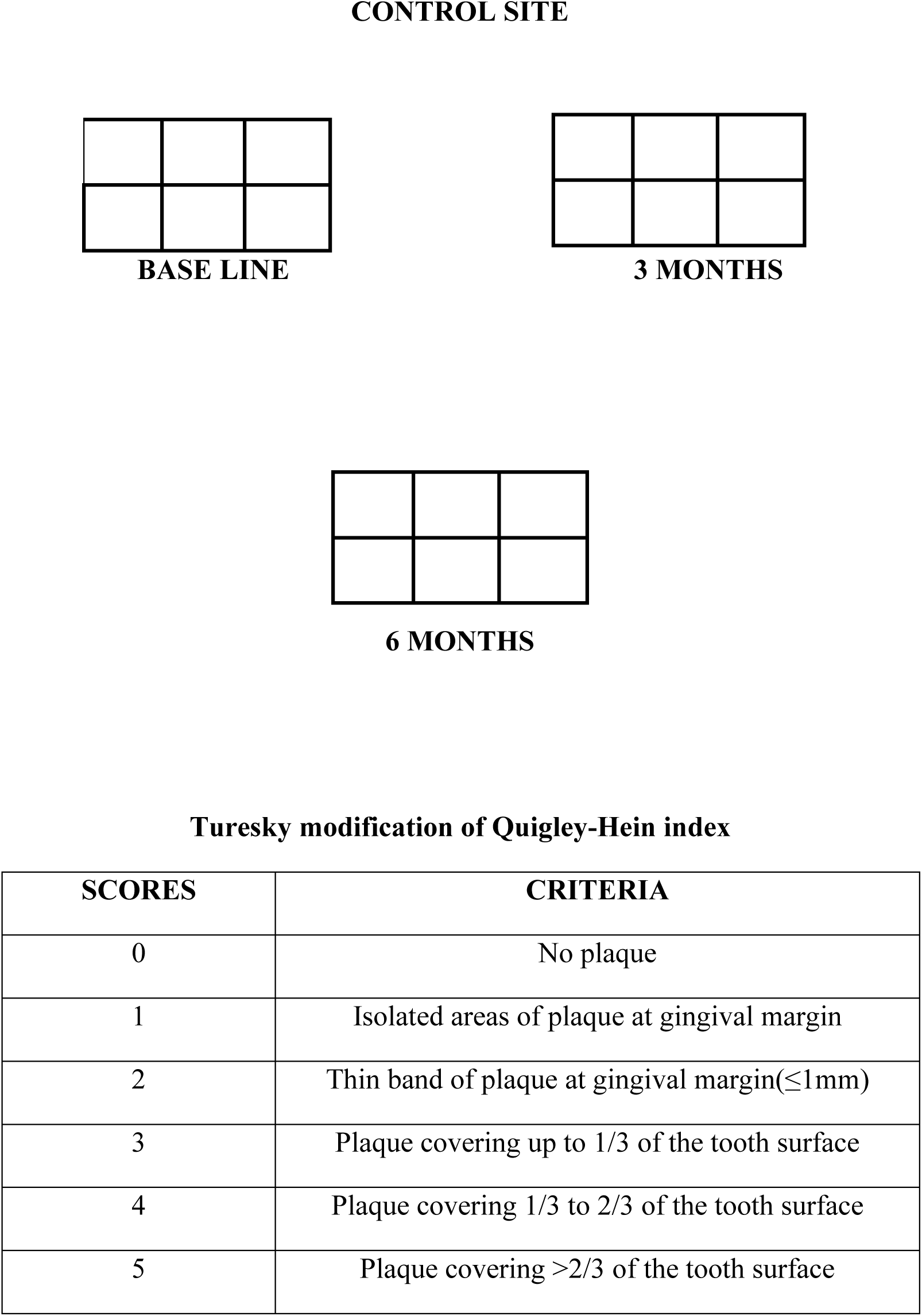

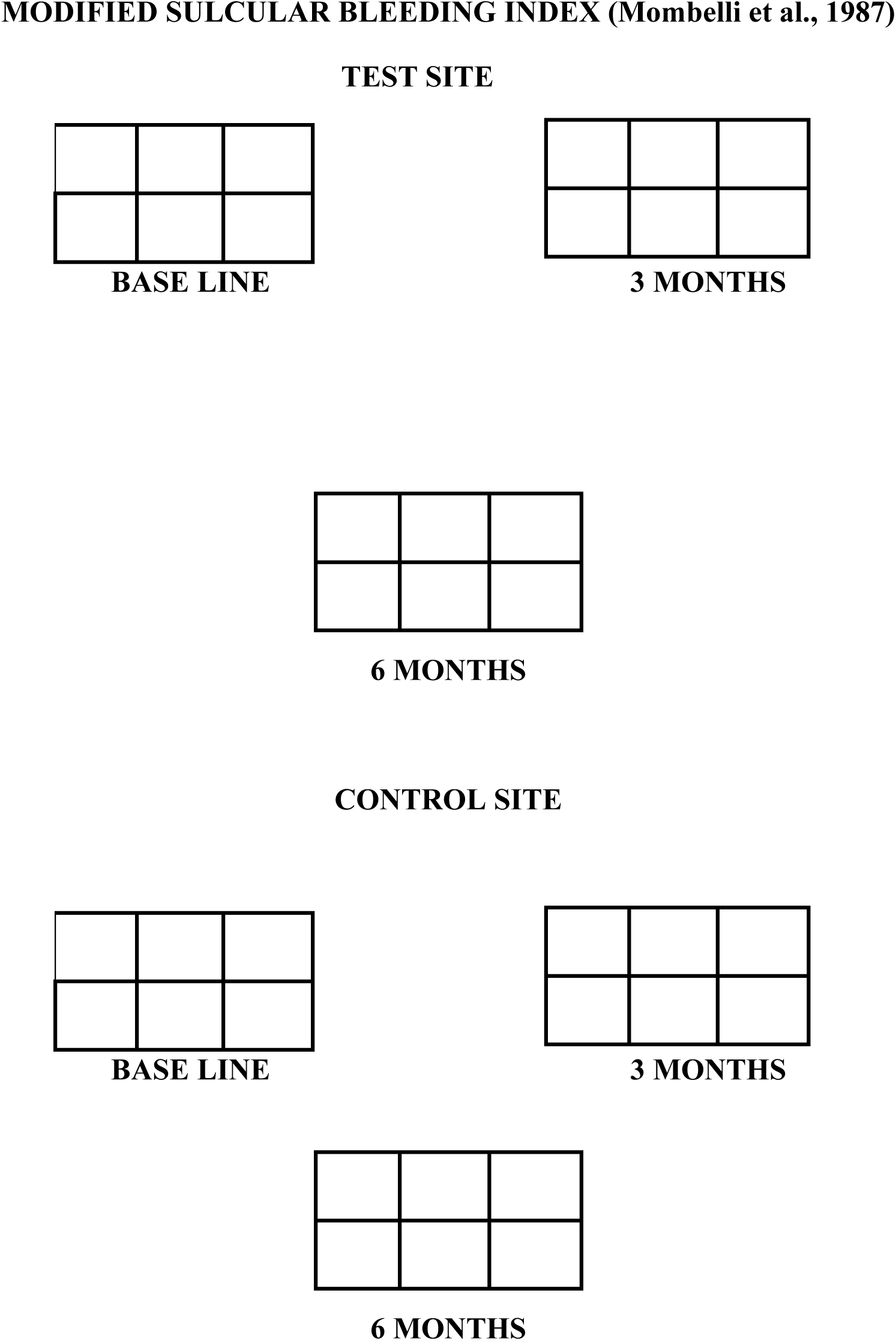

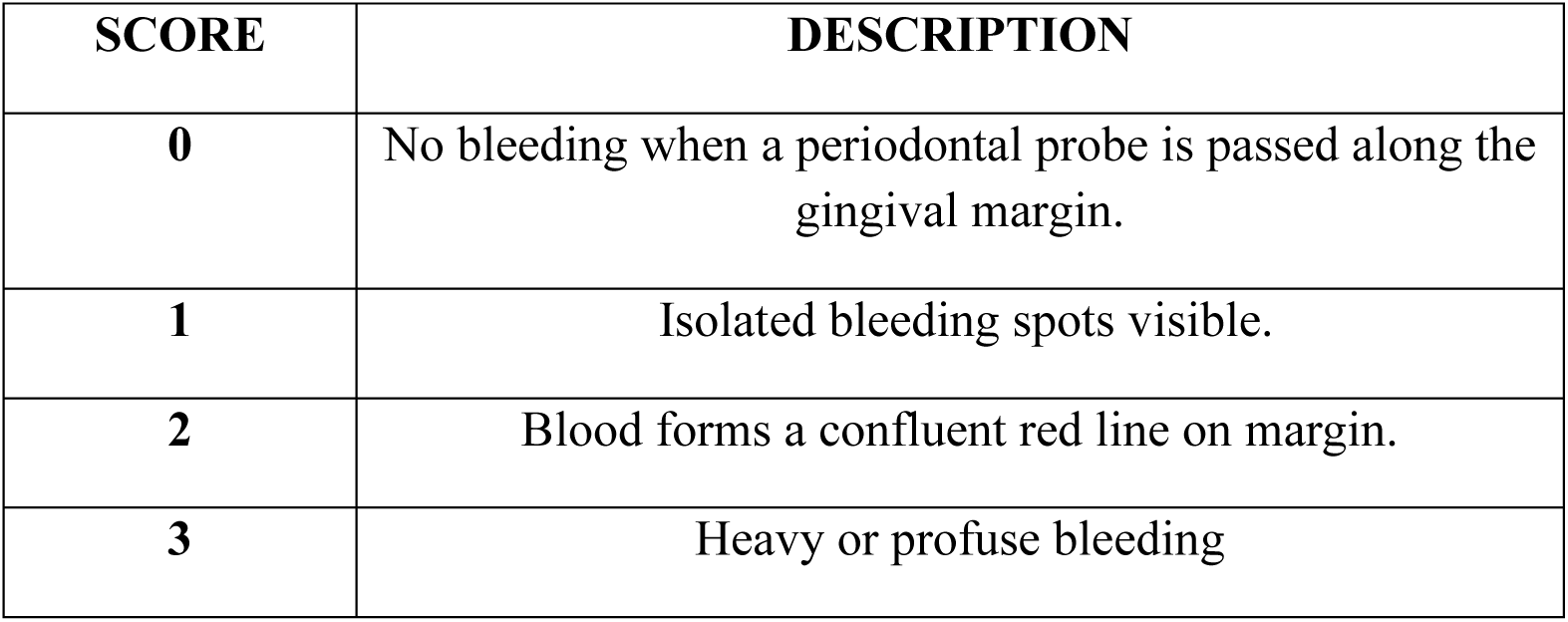

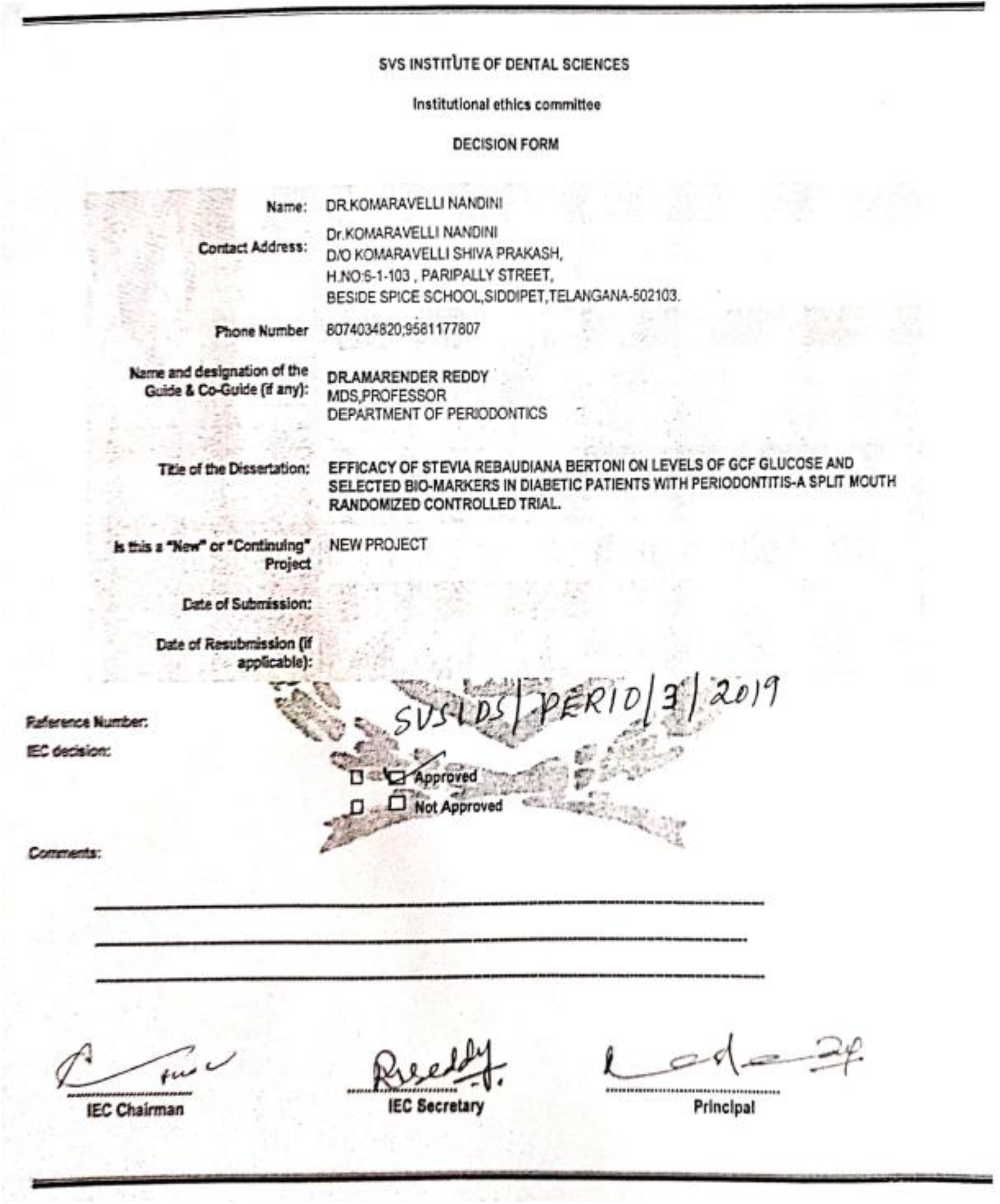

## References

1. Newman MG, Carranza FA, Takei H, Klokkevold PR. Carranzas clinical Periodontology. 10th ed. Elsevier health sciences; 2006.

2. Saini R, Marawar PP, Shete S, Saini S. Periodontitis, a true infection. J Glob Infect Dis. 2009; 1:149–50.

3. Mealey BL. Diabetes and periodontal disease: two sides of a coin. Compendium of continuing education in dentistry (Jamesburg, NJ: 1995). 2000; 21:943–6.

4. Grossi SG, Genco RJ. Periodontal disease and diabetes mellitus: a two-way relationship. Ann Periodontol. 1998; 3:51–61.

5. Soskolne WA. Epidemiological and clinical aspects of periodontal diseases in diabetics. Ann Periodontol. 1998; 3:3–12.

6. Iacopino AM, Cutler CW. Pathophysiological relationships between periodontitis and systemic disease: recent concepts involving serum lipids. J Periodontol. 2000; 71:1375–84.

7. Chang JC, Wu MC, Liu IM, Cheng JT. Increase of insulin sensitivity by stevioside in fructose-rich chow fed rats. Horm Metab Res 2005; 37:610–6.

8. Curi R, Alvarez M, Bazotte RB, Botion LM, Godoy JL, Bracht A. Effect of Stevia rebaudiana on glucose tolerance in normal adult humans. Braz J Med Biol Res 1986; 19:771–4.

9. Barriocanal LA, Palacios M, Benitez S, Jimenez JT, Jimenez N, et al. Apparent lack of pharmacological effect of steviol glycosides used as sweeteners in humans. A pilot study of repeated exposures in some normotensive and hypotensive individuals and in Type 1 and Type 2 diabetics. Regul Toxicol Pharmaco 2008; 51:37–41.

10. Contreras MS. Anticariogenic properties and effects on periodontal structures of Stevia rebaudiana Bertoni. Narrative review. J Oral Res. 2013; 2:158–66.

11. Vandana K, Reddy VC, Sudhir KM, Kumar K, Raju SH, Effectiveness of stevia as a mouth-rinse among 12–15-year-old schoolchildren in Nellore district, Andhra Pradesh-A randomized controlled trial. J Indian Soc Periodontol. 2017;21;37–43

12. Hanes PJ, Krishna R. Characteristics of inflammation common to both diabetes and periodontitis: Are predictive diagnosis and targeted preventive measures possible? EPMA J. 2010; 1:101–16.

13. Fitzsimmons TR, Sanders AE, Bartold PM, Slade GD. Local and systemic biomarkers in gingival crevicular fluid increase odds of periodontitis. J Clin Periodontol. 2010; 37:30–6.

14. Mohamed HG, Idris SB, Mustafa M, Ahmed MF, Ibrahim SO. Impact of Chronic Periodontitis on Levels of Glucoregulatory Biomarkers in Gingival Crevicular Fluid of Adults with and without Type 2 Diabetes. PLoS One. 2015;10: e0127660.

15. Jentsch HFR, Arnold N, Richter V, Deschner J. Salivary, gingival crevicular fluid and serum levels of ghrelin and chemerin in patients with periodontitis and overweight. J Periodontal Res. 2017; 52:1050–7.

16. Lalla E, Papapanou P. Diabetes mellitus and periodontitis: a tale of two common interrelated diseases. Nat Rev Endocrinol 7, 738–748;2011.

17. Preshaw PM, Alba AL, Herrera D, Jepsen S, Taylor R. Periodontitis and diabetes: a two-way relationship. Diabetologia. 2012; 55:21–31.

18. Taylor GW, Burt BA, Becker MP, Genco RJ, Shlossman M. Glycemic control and alveolar bone loss progression in type 2 diabetes. Ann Periodontol. 1998; 3:30–9.

19. Chiu SY, Lai H, Yen AM, Fann JC, Chen HH. Temporal sequence of the bidirectional relationship between hyperglycemia and periodontal disease: a community-based study of 5,885 Taiwanese aged 35-44 years (KCIS No. 32). Acta Diabetol. 2015; 52:123–31.

20. Jimenez M, Hu FB, Marino M, Li Y, Joshipura KJ. Type 2 diabetes mellitus and 20-year incidence of periodontitis and tooth loss. Diabetes Res Clin Pract. 2012; 98:494–500.

21. Morita I, Inagaki K, Nakamura F, Mizuno K, Sabbah W. Relationship between periodontal status and levels of glycated hemoglobin. J Dent Res. 2012; 91:161–6.

22. Nelson RG, Shlossman M, Budding LM, Saad MF, Genco RJ, Knowler WC. Periodontal disease and NIDDM in Pima Indians. Diabetes Care. 1990; 13:836–40.

23. McAllister BS, Haghighat K. Bone augmentation techniques. J Periodontol. 2007; 78:377–96.

24. Samuel P, Ayoob KT, Jeppesen PB, Rogers PJ, Mathews R. Stevia Leaf to Stevia Sweetener: Exploring Its Science, Benefits, and Future Potential. J Nutr. 2018; 148:1186S–1205S.

25. Farhat G, Berset V, Moore L. Effects of Stevia Extract on Postprandial Glucose Response, Satiety and Energy Intake: A Three-Arm Crossover Trial. Nutrients. 2019; 11:30–6.

26. Goyal SK, Samsher, Goyal RK. Stevia (Stevia rebaudiana) a bio-sweetener: a review. Int J Food Sci Nutr. 2010; 61:1–10.

27. Rojas E, Bermudez V, Motlaghzadeh Y, Contreras J, Mantilla LP. Stevia rebaudiana Bertoni and Its Effects in Human Disease: Emphasizing Its Role in Inflammation, Atherosclerosis and Metabolic Syndrome. Curr Nutr Rep. 2018.

28. Carrera-Lanestosa A, Moguel-Ordonez Y, Segura-Campos M. Stevia rebaudiana Bertoni: A Natural Alternative for Treating Diseases Associated with Metabolic Syndrome. J Med Food. 2017; 20:933–43.

29. Shivanna N, Naika M, Khanum F, Kaul VK. Antioxidant, anti-diabetic and renal protective properties of Stevia rebaudiana. J Diabetes Complications. 2013; 27:103–13.

30. Akbarzadeh S, Eskandari F, Bargahi A, Bazzi P, Daneshi A. The effect of Stevia rebaudiana on serum omentin and visfatin level in STZ-induced diabetic rats. J Diet Suppl. 2015; 12:11–22.

31. Ilic V, Vukmirovic S, Stilinovic N, Capo I, Arsenovic M. Insight into anti-diabetic effect of low dose of stevioside. Biomed Pharmacother. 2017; 90:216–21.

32. Gregersen S, Jeppesen PB, Holst JJ, Hermansen K. Antihyperglycemic effects of stevioside in type 2 diabetic subjects. Metabolism. 2004; 53:73–6.

33. Ajami M, Seyfi M, Pouri Hosseini F, Naseri P. Effects of stevia on glycemic and lipid profile of type 2 diabetic patients: A randomized controlled trial. Avicenna J Phytomed. 2020; 10:118–27.

34. Jan SA, Habib N, Shinwari ZK, Ali M, Ali N. The anti-diabetic activities of natural sweetener plant Stevia: an updated review. SN Applied Sciences. 2021; 3:1–6.

35. Ray J, Kumar S, Laor D, Soni L, McFarlane SI. Effects of Stevia Rebaudiana on Glucose Homeostasis, Blood Pressure and Inflammation: A Critical Review of Past and Current Research Evidence. International journal of clinical research & trials. 2020;5.

36. Subramaniam R, Mittal S, Hiregoudar M, Ratan GN, Chandu GN. Antimicrobial activity of Stevioside on periodontal pathogens and Candida Albicans-An invitro study. Journal of Indian Association of Public Health Dentistry. 2011; 9:325.

37. Boonkaewwan C, Toskulkao C, Vongsakul M. Anti-inflammatory and immunomodulatory activities of stevioside and its metabolite steviol on THP-1 cells. J Agri Food Chem. 2006; 54:785–9.

38. Iacopino AM. Periodontitis and diabetes interrelationships: role of inflammation. Annals of periodontology. 2001; 6:125–37.

39. Chang PC, Lim LP. Interrelationships of periodontitis and diabetes: A review of the current literature. Journal of Dental Sciences. 2012; 7:272–82.

40. Yilmaz G, Kirzioglu FY, Doguc DK, Kocak H, Orhan H. Ghrelin levels in chronic periodontitis patients. Odontology. 2014; 102:59–67.

41. Patnaik K, Pradeep AR, Nagpal K, Karvekar S, Raju A. Human chemerin correlation in gingival crevicular fluid and tear fluid as markers of inflammation in chronic periodontitis and type-2 diabetes mellitus. J InvestigClin Dent. 2017;8.

42. Dogan SB, Balli U, Dede FO, Sertoglu E, Tazegul K. Chemerin as a Novel Crevicular Fluid Marker of Patients with Periodontitis and Type 2 Diabetes Mellitus. J Periodontol. 2016; 87:923–33.

43. Devanoorkar A, Kathariya R, Guttiganur N, Gopalakrishnan D, Bagchi P. Resistin: a potential biomarker for periodontitis influenced diabetes mellitus and diabetes induced periodontitis. Disease markers. 2014.

44. Yilmaz D, Caglayan F, Buber E, Kononen E, Guncu GN. Gingival crevicular fluid levels of human beta-defensin-1 in type 2 diabetes mellitus and periodontitis. Clinical oral investigations. 2018; 22:2135–40.

45. Joshi A, Maddipati S, Chatterjee A, Lihala R, Gupta A. Gingival crevicular fluid resistin levels in chronic periodontitis with type 2 diabetes before and after non-surgical periodontal therapy: A clinico-biochemical study. Indian J Dent Res. 2019; 30:47–51.

46. Ahuja CR, Kolte AP, Kolte RA, Gupta M, Chari S. Effect of non-surgical periodontal treatment on gingival crevicular fluid and serum leptin levels in periodontally healthy chronic periodontitis and chronic periodontitis patients with type 2 diabetes mellitus. Journal of investigative and clinical dentistry. 2019;10: e12420.

47. Bajaj D, Dhadse D, Prasad V, Baliga D, Jaiswal D. Assessment and Comparison of Ghrelin and Chemerin Levels in Gingival Crevicular Fluid and Serum as Predictive Biomarkers in Aggressive Periodontitis Patients: A Study Protocol. European Journal of Molecular & Clinical Medicine. 2020; 7:2027–33.

48. Bozkurt Dogan S, Ongoz Dede F, Balli U, Sertoglu E. Levels of vaspin and omentin-1 in gingival crevicular fluid as potential markers of inflammation in patients with chronic periodontitis and type 2 diabetes mellitus. J Oral Sci. 2016; 58:379–89.

49. Barros SP, Williams R, Offenbacher S, Morelli T. Gingival crevicular fluid as a source of biomarkers for periodontitis. Periodontol 2000. 2016; 70:53–64.

50. Fatima T, Khurshid Z, Rehman A, Srivastava KC, Shrivastava D. Gingival Crevicular Fluid (GCF): A Diagnostic Tool for the Detection of Periodontal Health and Diseases. Molecules. 2021; 26:1208.

51. Nagarathna R, Bali P, Anand A, Srivastava V, Nagendra HR. Prevalence of Diabetes and Its Determinants in the Young Adults Indian Population-Call for Yoga Intervention. Frontiers in endocrinology. 2020;11.

52. Kumar R, Singh V. Cultivation of Stevia [Stevia rebaudiana (Bert.) Bertoni]: A Comprehensive Review. Adv Agron 2006; 89: 137–77.

53. Cioni P, Morelli I, Andolfi L, Macchia M, Ceccarini L. Qualitative and quantitative analysis of essential oils of five lines Stevia rebaudiana Bert. Genotypes cultivated in Pisa (Italy). J Essent Oil Res 2006; 18: 76–9.

54. Totte N, Ende W, Van Damme E, Geuns J. Cloning and heterologous expression of early genes in gibberellin and Steviol biosynthesis via the methylerythritol phosphate pathway in Stevia rebaudiana. Can J Bot 2003; 81: 517–22.

55. Bender C, Graziano S, Zimmermann BF. Study of Stevia rebaudiana Bertoni antioxidant activities and cellular properties. Int J Food Sci Nutr. 2015; 66:553–8.

56. Riera CE, Vogel H, Simon SA, le Coutre J. Artificial sweeteners and salts producing a metallic taste sensation activate TRPV1 receptors. Am J Physiol Regul Integr Comp Physiol. 2007;293: R626–34.

57. Tarka S, Roberts A. Stevia: It’s Not Just About Calories. Open Obes J 2010; 2:101–9

58. Melis M, Rocha S, Augusto A. Steviol effect, a glycoside of Stevia rebaudiana, on glucose clearances in rats. Braz J Biol 2009; 69: 371–4.

59. Curi R, Alvarez M, Bazotte RB. Effect of Stevia rebaudiana on glucose tolerance in normal adult humans. Braz J Med Biol Res 1986; 19: 771–4.

60. Krentz AJ, Hompesch M. Glucose: archetypal biomarker in diabetes diagnosis, clinical management and research. Biomarkers in medicine. 2016; 10:1153–66.

61. Pontes Andersen CC, Flyvbjerg A, Buschard K, Holmstrup P. Relationship between periodontitis and diabetes: lessons from rodent studies. Journal of periodontology. 2007; 78:1264–75.

62. Groschl M, Topf HG, Bohlender J, Zenk J, Rauh M. Identification of ghrelin in human saliva: production by the salivary glands and potential role in proliferation of oral keratinocytes. Clinical chemistry. 2005; 51:997–1006.

63. Allen EM, Chapple IL. The Relationship Between Periodontitis and Glycaemic Control in Type 2 Diabetes. Eur Endocrinol. 2012; 8:89–93.

64. Teshome A, Yitayeh A. The effect of periodontal therapy on glycaemic control and fasting plasma glucose level in type 2 diabetic patients: systematic review and meta-analysis. BMC oral health. 2017; 17:1–1.

65. Mauri-Obradors E, Merlos A, Vinas M. Benefits of non-surgical periodontal treatment in patients with type 2 diabetes mellitus and chronic periodontitis: A randomized controlled trial. J Clin Periodontol. 2018; 45:345–53.

66. Munjal A, Jain Y, Kote S, Krishnan V, Passi D. A study on the change in HbA1c levels before and after non-surgical periodontal therapy in type-2 diabetes mellitus in generalized periodontitis. J Family Med Prim Care. 2019; 8:1326–9.

67. Mizuno H, Ekuni D, Maruyama T, Wada J, Morita M. The effects of non-surgical periodontal treatment on glycaemic control, oxidative stress balance and quality of life in patients with type 2 diabetes: A randomized clinical trial. PLoS One. 2017;12: e0188171.

68. Hsu YT, Nair M, Angelov N, Lalla E, Lee CT. Impact of diabetes on clinical periodontal outcomes following non-surgical periodontal therapy. J Clin Periodontol. 2019; 46:206–17.

69. Kujur RS, Singh V, Ram M, Yadava HN, Roy BK. Antidiabetic activity and phytochemical screening of crude extract of Stevia rebaudiana in alloxan-induced diabetic rats. Pharmacognosy research. 2010; 2:258.

70. Saravanan, R.; Ramachandran, V. Modulating efficacy of Rebaudioside A, a diterpenoid on antioxidant and circulatory lipids in experimental diabetic rats. Environ. Toxicol. Pharmacol. 2013; 36:472–83.

71. Chang JC, Wu MC, Liu IM, Cheng JT. Increase of insulin sensitivity by stevioside in fructose-rich chow-fed rats. Horm Metab Res. 2005; 37:610–16.

72. Anton SD, Martin CK, Han H, Coulon S, Williamson DA. Effects of stevia, aspartame, and sucrose on food intake, satiety, and postprandial glucose and insulin levels. Appetite. 2010; 55:37–43.

73. Sharma R, Yadav R, Manivannan Ozbayer C, Degirmenci I, Hulyam K. Study of effect of Stevia rebaudiana bertoni on oxidative stress in type-2 diabetic rat models. Biomed Aging Pathol 2012; 2:126–31

74. Jeppenson PB, Rolfsen SE, Colombo M, Agger A et al., Antihyperglycemic and blood pressure reducing effects of stevioside in the diabetic goto-kakizaki rat. Metabolism. 2003; 52:372–8.

75. Ritu M, Nandini J. Nutritional composition of Stevia rebaudiana, a sweet herb, and its hypoglycaemic and hypolipidemic effect on patients with non-insulin dependent diabetes mellitus. Journal of the Science of Food and Agriculture. 2016; 96:4231–4.

76. Abundula R, Matchkov VV, Nilsson H, Hermansen K. Rebaudioside A directly stimulates insulin secretion from pancreatic beta cells: A glucose dependent action via inhibition of ATP sensitive K+ channels. Diabetes, Obes Metab. 2008; 10:1074–85.

77. Cummings DE, Purnell JQ, Frayo RS, Schmidova K, Weigle DS. A pre-prandial rise in plasma ghrelin levels suggests a role in meal initiation in humans. Diabetes. 2001; 50:1714–9.

78. Flanagan DE, Evans ML, Heptulla RA, Tamborlane WV, Sherwin RS. The influence of insulin on circulating ghrelin. American Journal of Physiology-Endocrinology and Metabolism. 2003;284: E313–6.

79. Tschop M, Bidlingmaier M, Landgraf R, Folwaczny C. Post-prandial decrease of circulating human ghrelin levels. Journal of endocrinological investigation. 2001;24:RC19–21.

80. Poher AL, Tschöp MH, Müller TD. Ghrelin regulation of glucose metabolism. Peptides. 2018; 100:236–42.

81. Banks KA, Murphy KG. Role of ghrelin in glucose homeostasis and diabetes. Diabetes Management. 2013; 3:171.

82. Pöykkö SM, Kellokoski E, Hörkkö S, Ukkola O. Low plasma ghrelin is associated with insulin resistance, hypertension, and the prevalence of type 2 diabetes. Diabetes. 2003; 52:2546–53.

83. Tiwari BS, Ankola AV, Sankeshwari RM, Patil P, Kashyap BR. Comparison of effectiveness for Stevia rebaudiana and chlorhexidine mouth rinses on plaque and gingival scores among 12–15-year-old government school children in Belagavi City – A randomized controlled trail. Indian J Health Sci Biomed Res 2020; 13:32–6.

84. De Slavutzky SM. Stevia and sucrose effect on plaque formation. J Fur Verbraucherschutz Leben 2010; 5:213–6.

85. Lin L H, Lee L, Sheu S, Lin P. Study on the stevioside analogues of stevioside, steviol, and isosteviol 19-alkyl amide dimers: synthesis and cytotoxic and antibacterial activity. Chem Pharm Bull 2004; 52:1117–22.

86. Bolanos J, Ramirez J. Effects of Stevia rebaudiana bertoni extract on periodontal disease. En: XXIII scientific dental congress ACCO; 2007.

87. Stengel A, Goebel M, Wang L, Lambrecht NW. Lipopolysaccharide differentially decreases plasma acyl and desacyl ghrelin levels in rats: potential role of the circulating ghrelin-acylating enzyme GOAT. Peptides. 2010; 31:1689–96.

88. Wang L, Basa NR, Shaikh A, et al. LPS inhibits fasted plasma ghrelin levels in rats: role of IL-1 and PGs and functional implications. Am J Physiol Gastrointest Liver Physiol. 2006;291: G611–20.

89. Huang CX, Yuan MJ, Huang H, et al. Ghrelin inhibits post-infarct myocardial remodelling and improves cardiac function through antiinflammation effect. Peptides. 2009; 30:2286–91.

90. Au CC, Docanto MM, Zahid H, et al. Des-acyl ghrelin inhibits the capacity of macrophages to stimulate the expression of aromatase in breast adipose stromal cells. J Steroid Biochem Mol Biol. 2017; 170:49–53.

91. Waseem T, Duxbury M, Ito H, Robinson MK. Exogenous ghrelin modulates release of pro-inflammatory and anti-inflammatory cytokines in LPS-stimulated macrophages through distinct signalling pathways. Surgery. 2008; 143:334–42.

92. Warzecha Z, Kownacki P, Ceranowicz P, Dembinski M, Cieszkowski J, Dembinski A. Ghrelin accelerates the healing of oral ulcers in nonsialoadenectomized and sialoadenectomized rats. J Physiol Pharmacol. 2013; 64:657–68.

93. Akki R, Raghay K, Errami M. Potentiality of ghrelin as antioxidant and protective agent. Redox Report. 2021; 26:71–9.

94. Almehmadi AH, Alghamdi F. Biomarkers of alveolar bone resorption in gingival crevicular fluid: A systematic review. Archives of oral biology. 2018; 93:12–21.

95. Kc S, Wang XZ, Gallagher JE. Diagnostic sensitivity and specificity of host-derived salivary biomarkers in periodontal disease amongst adults: systematic review. Journal of clinical periodontology. 2020; 47:289–308.

96. Aziz AS, Kalekar MG, Benjamin T, Bijle MN. Effect of nonsurgical periodontal therapy on some oxidative stress markers in patients with chronic periodontitis: A biochemical study. World J Dent. 2013; 4:17–23.

97. Ruiz JC, Ordoñez YB, Basto ÁM, Campos MR. Antioxidant capacity of leaf extracts from two Stevia rebaudiana Bertoni varieties adapted to cultivation in Mexico. Nutricion hospitalaria. 2015; 31:1163–70.

98. Ameer K, Jiang GH, Amir RM, Eun JB. Antioxidant potential of Stevia rebaudiana (Bertoni). InPathology 2020 (pp. 345-356). Academic Press.

